# A Systematic Review of the Quantitative markers of speech and language of the Frontotemporal Degeneration Spectrum and their potential for cross-linguistic implementation

**DOI:** 10.1101/2024.01.05.24300888

**Authors:** Rosie Coppieters, Arabella Bouzigues, Lize Jiskoot, Maxime Montembeault, Boon Lead Tee, Genetic Frontotemporal dementia Initiative (GENFI), Jonathan D. Rohrer, Rose Bruffaerts

## Abstract

Frontotemporal dementia (FTD) is a neurodegenerative disease spectrum with an urgent need for reliable biomarkers for early diagnosis and monitoring. Speech and language changes occur in the early stages of FTD and offer a potential non-invasive, early, and accessible diagnostic tool. The use of speech and language markers in this disease spectrum is limited by the fact that most studies investigate English-speaking patients. This systematic review examines the literature on psychoacoustic and linguistic features of speech that occur across the FTD spectrum across as many different languages as possible. 76 papers were identified that investigate psychoacoustic and linguistic markers in discursive speech. 75% of these papers studied English-speaking patients. The most generalisable features found across different languages, are speech rate, articulation rate, pause frequency, total pause duration, noun: verb ratio, and total number of nouns. While there are clear interlinguistic differences across patient groups, the results show promise for implementation of cross-linguistic markers of speech and language across the FTD spectrum, particularly for psychoacoustic features.

## 1. INTRODUCTION

Frontotemporal dementia (FTD) gives rise to a spectrum of clinical phenotypes with variable degrees of speech and language pathology (Moore et al., 2020; Rohrer et al., 2015). All variants of primary progressive aphasia (PPA) by definition present with prominent abnormalities of speech and/or language (Gorno-Tempini et al., 2011), but the FTD spectrum as a whole is strongly associated with such impairments. In behavioral variant FTD (bvFTD), word retrieval, comprehension, reading, writing, verbal and non-verbal semantic knowledge, as well as prosody of speech are impaired, while motor speech and repetition abilities remain generally conserved (Geraudie et al., 2021; Samra et al., 2023). On the other side of the spectrum, progressive supranuclear palsy (PSP) and corticobasal syndrome (CBS) have been more often associated with motor speech impairments such as apraxia of speech and dysarthria, though more recent research also shows impairment in language abilities, such as confrontation naming, fluency, sentence comprehension and production (Peterson et al., 2019). Meanwhile, FTD combined with amyotrophic lateral sclerosis (FTD-ALS) can occasionally present with agrammatism and/or apraxia of speech, as well as comprehension deficits on the single word and sentence level (Rusina et al., 2021).

While the first clinical trials for genetic FTD are being rolled out (Boeve et al., 2022), there is an unmet clinical need for reliable non-invasive markers to monitor disease progression and therapeutic effects. Quantitative analysis of speech and language in FTD could potentially provide objective, low-cost and sensitive markers suitable for this purpose. Specifically, psychoacoustic markers can be used to quantify motor speech disorders such as apraxia of speech and dysarthria, while linguistic markers may quantify single word use or detect sentence construction abnormalities.

One of the important challenges regarding the clinical implementation of psychoacoustic and linguistic markers is to identify which, if any, markers are applicable across different languages. There is a paucity of published research on the speech and language changes that occur in FTD spectrum disorders in non-English speaking patients (García et al., 2023), therefore, it would be beneficial to determine which changes in speech and language are generalizable across languages and which are language specific. Research has shown significant interlinguistic differences between patients across the FTD spectrum (García et al., 2023). For instance, significant differences have been found between English and Italian patients with nfvPPA in motor speech and syntactic complexity (Canu et al., 2020).

There are numerous approaches to investigating the changes that occur in speech and language. Connected speech (comprising of consecutive words forming utterances) offers a wealth of information about the cognitive state of a patient, and countless variables can be extracted from a relatively small sample of connected speech. The cookie theft picture description task for instance, one of the most commonly used tasks to generate connected speech (Goodglass & Kaplan, 1972), takes less than two minutes to administer and can be carried out by experts and non-experts alike. There are also numerous alternatives to picture description tasks, for instance, semi-structured interviews (Knibb et al., 2009), narrative tasks (Ash et al., 2006), reading tasks (Baque et al., 2022), and repetition tasks (Bouvier et al., 2021). Another advantage of picture description tasks such as the cookie theft picture description task is that large existing databases of speech samples from patients with FTD as well as from controls can be used as comparisons for newly collected data. These large databases will be beneficial in creating machine learning models to diagnose FTD using (semi) automated analysis of speech.

In this paper, we performed a systematic review of the known quantitative psychoacoustic and linguistic markers in connected speech across the FTD spectrum to inform future cross-linguistic research and clinical implementation.

## 2. METHODS

A systematic review of the literature on the quantitative psychoacoustic and linguistic markers of FTD was carried out to determine the most relevant features of speech and language to analyze in discursive speech samples, and to determine which features were the most generalizable across different languages. Using the databases SCOPUS and Web of Science the relevant literature was identified and screened on the 1^st^ of March 2023. The search terms used were “FTLD”, “primary progressive aphasia”, “frontotemporal dementia”, “semantic dementia”, “non-fluent primary progressive aphasia”, “progressive supranuclear palsy”, “corticobasal degeneration”, “ALSFTD”, “speech”, and “language”. The disorders were grouped by an ‘OR’ operator and combined with the speech and language search terms with the ‘AND’ operator, which were also grouped with the ‘OR’ operator. All words are MeSH terms.

This search yielded 5272 results in total including duplicates, which were manually removed, leaving 3771 peer-reviewed papers (see Figure 1). Using Rayyan (Ouzzani et al., 2016), the titles and abstracts were triaged to filter for the relevant papers. Papers were included if they studied a population of patients with frontotemporal lobar degeneration and obtained results relating to the psychoacoustic or linguistic properties of connected speech or discourse of the patients. Patients with non-fluent variant PPA (nfvPPA), primary progressive apraxia of speech (PPAOS), logopenic variant PPA (lvPPA), semantic variant PPA (svPPA), semantic dementia (SD), mixed PPA (mxPPA), bvFTD, CBS, PSP, and ALS-FTD were included. Only patient groups that were diagnosed according to international consensus criteria were included. Papers were excluded if there was no control group, or if they were case studies or review papers.

**Figure 1.**
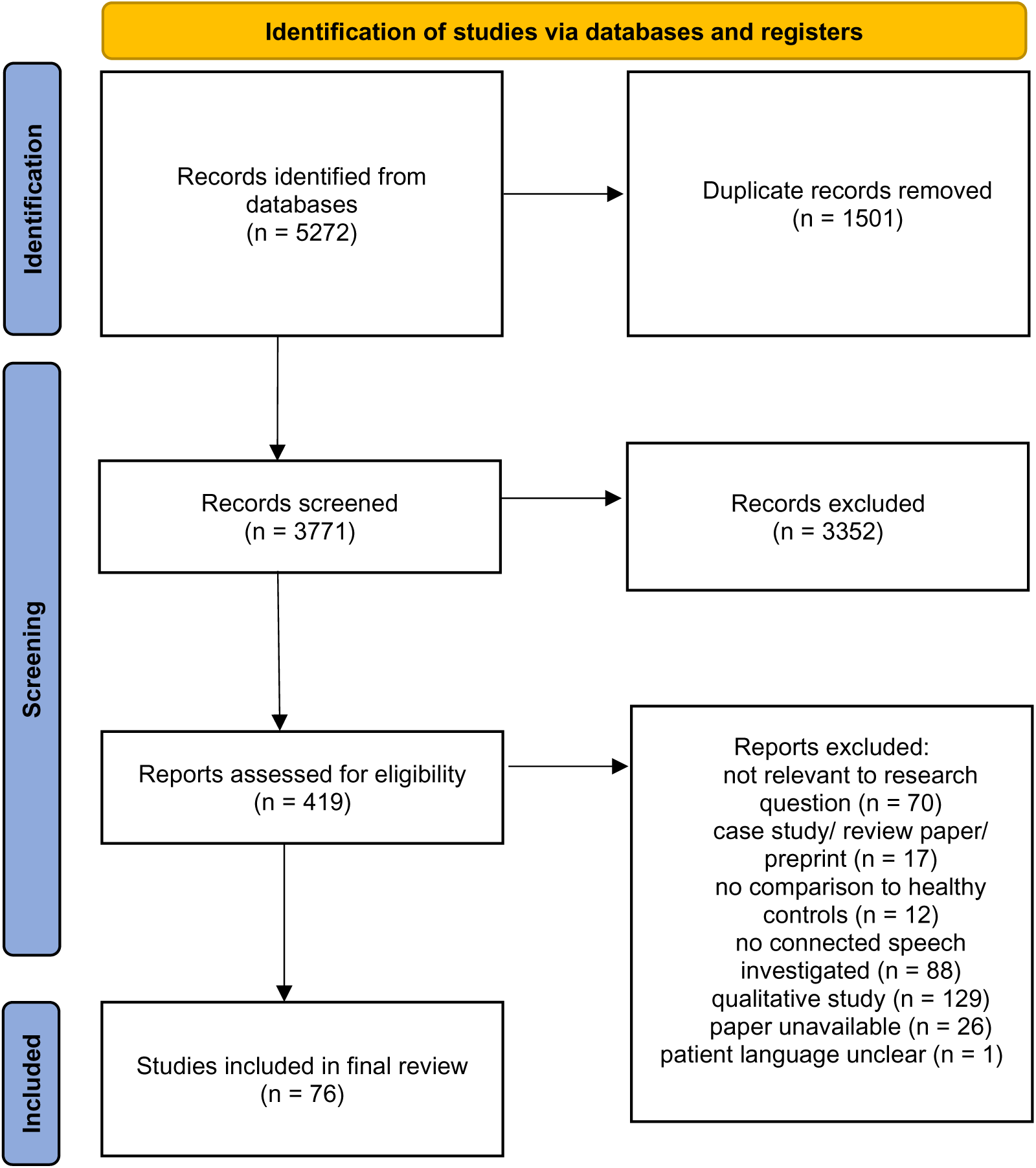
PRISMA flowchart of methods of systematic literature review.

With these inclusion and exclusion criteria, a total of 419 papers were selected for further reading. The following data was extracted from these 419 papers: (i) the sample size, (ii) the language of the patients, (iii) the methods used to obtain the speech and language features, (iv) the psychoacoustic and linguistic features investigated, and (v) the relevant findings.

Papers published in all languages were considered. Five included papers were only available in Spanish, one only available in German, the rest were written in English (though not all about English-speaking patients).

Upon further reading and data extraction, 76 papers were selected for inclusion in the final paper. The date range of these papers was from May 1997 to October 2022.

Based on the 76 papers, there were a total of 342 features of speech compared in the patient groups and healthy controls in discourse. 44 of these features are studied in more than one language.

In some papers the PPA patients were categorized as “mixed” or “undefined”. The results that were relevant to this review were considered separately and then the “mixed” PPA group was combined with the nfvPPA category when comparing quantitative values.

## 3. RESULTS

76 papers studying discourse in patients with FTD were included in the final review. 57 of these papers (75% of all papers) studied English-speaking patients, the remaining 19 papers studied patients speaking Spanish (Baque et al., 2022; Matias-Guiu et al., 2020, 2022), Czech (Daoudi et al., 2022; Rusz et al., 2015; Skrabal et al., 2020), Italian (Catricala et al., 2019; Silveri et al., 2014), French (Bouvier et al., 2021; Macoir et al., 2021), German (Hohlbaum et al., 2018; Staiger A., 2017), Dutch (Bruffaerts et al., 2022), Greek (Karpathiou & Kambanaros, 2022; Koukoulioti et al., 2018, 2020; Potagas et al., 2022), Hindi (Sachin et al., 2008), and Korean (Suh et al., 2010) (Figure 2a). Figure 2b shows the geographical representation of the published papers, with a paucity of languages from South America, Asia, and Africa. As Figure 2c shows, the language distribution of the literature is in no way representative of the total population of each language, with English being drastically overrepresented relative its total population of native speakers. The vast majority of these papers describe patients with sporadic FTD (99%).

**Figure 2.**
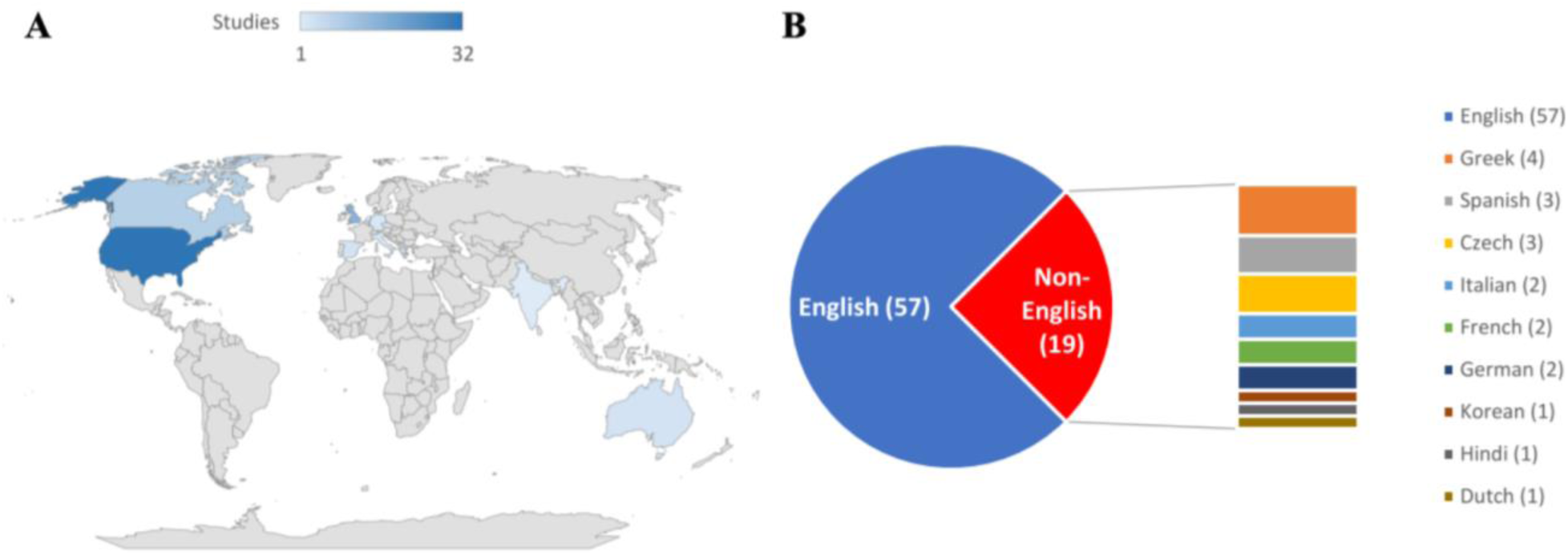
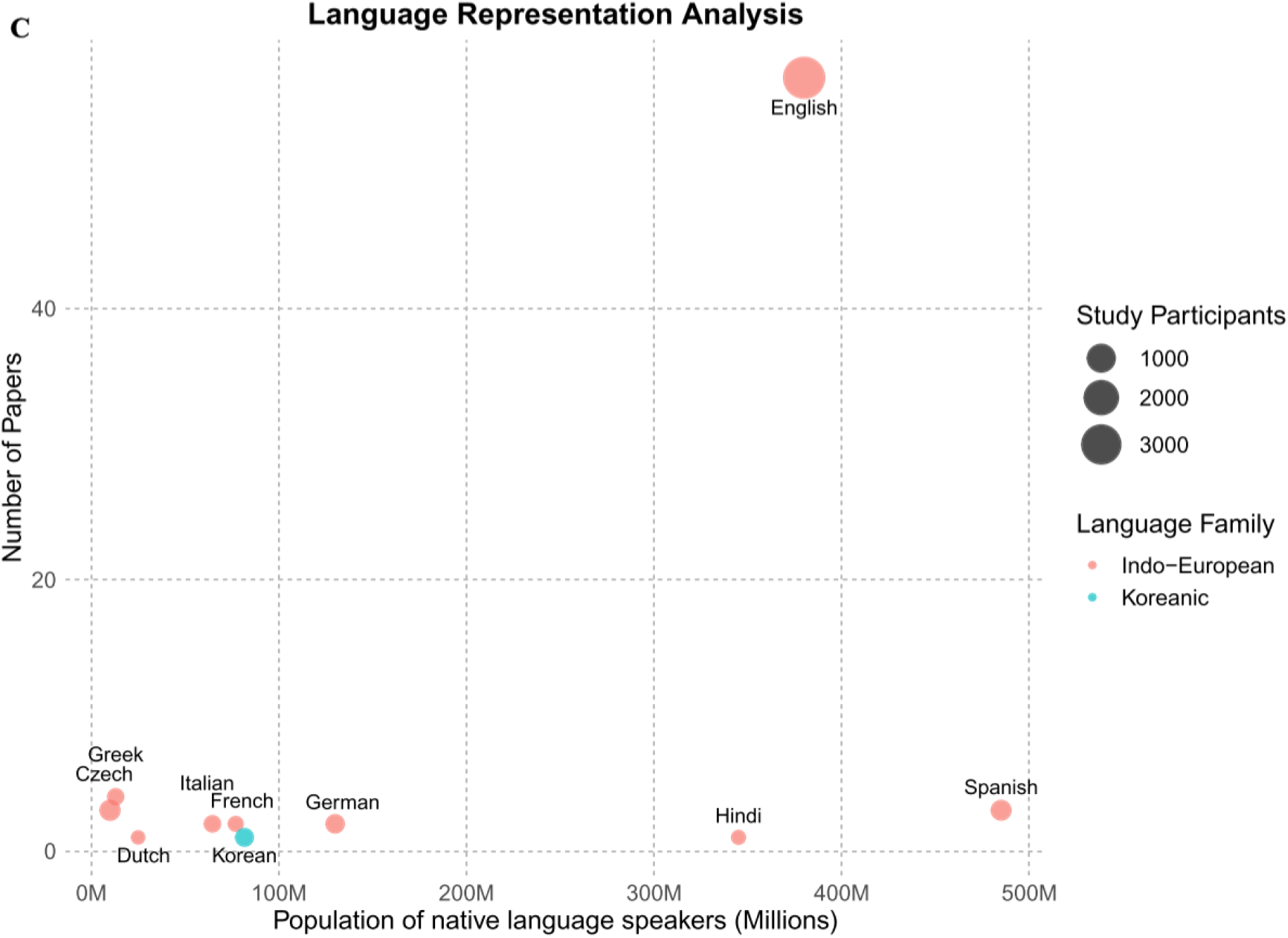
The (A) geographical locations, (B) languages, and (C) language representation analysis of the 76 included papers.

**Figure 3.**
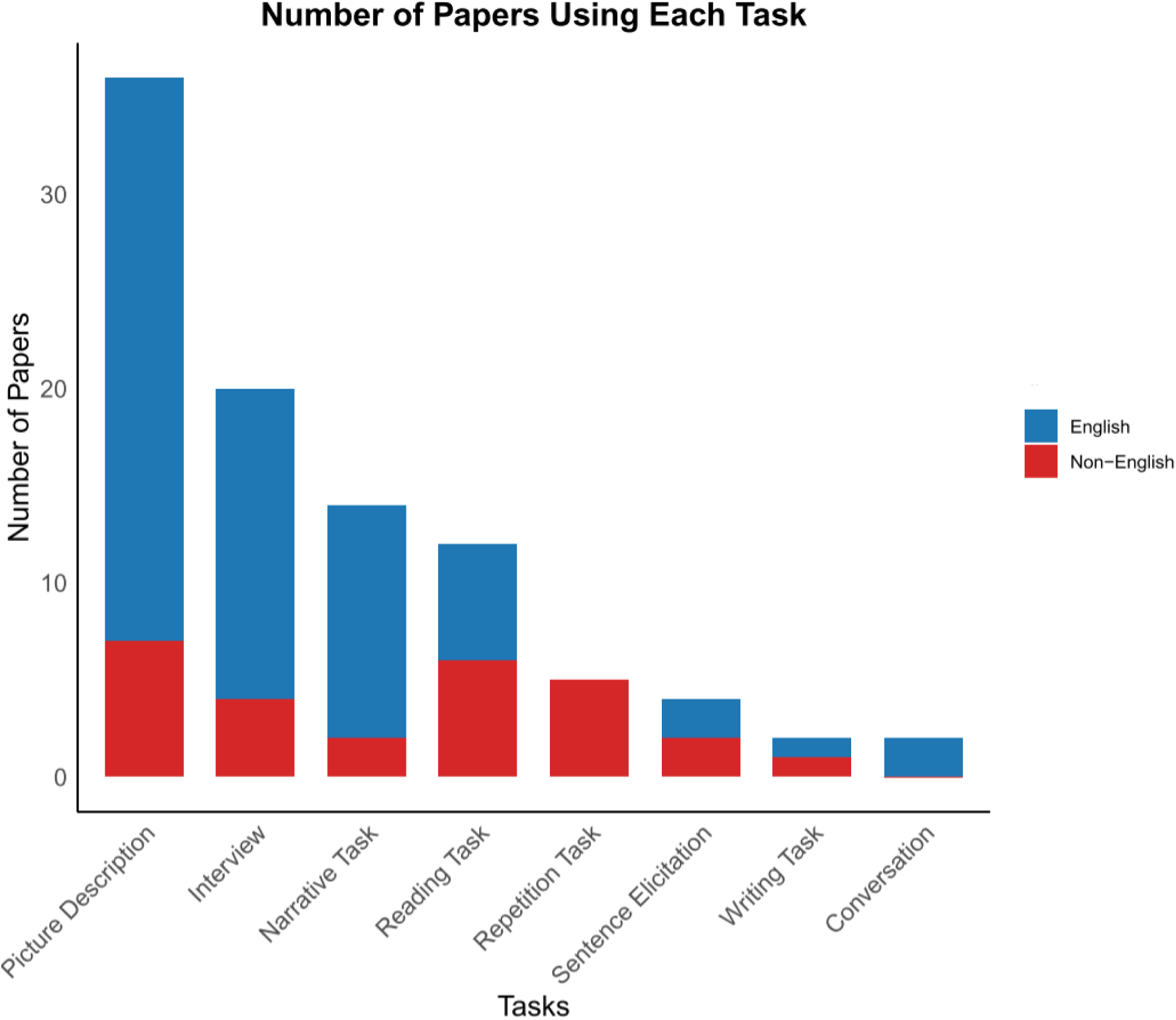
The tasks used by the papers in the review.

As previously stated, only papers using tasks which elicited connected speech were included. The tasks chosen were varied, but the most commonly used were picture description, interviews, narrative, and reading tasks. The cookie theft picture description task was the most widely used task, in a total of 23 papers with patients speaking English, Spanish, Dutch, Italian, and Greek. Sentence repetition tasks were only used in papers studying non-English-speaking patients. Conversely, conversation as a task was not used by any papers studying non-English-speaking patients, and was only used by two papers studying English-speaking patients.

The psychoacoustic and linguistic features of speech across the FTD spectrum were extracted, resulting in 342 features. The features were grouped based on the categorisation method of Boschi et. al. (2017) for connected speech in neurodegenerative disorders (2017), comprising of the following five categories: phonetic-phonological, lexico-semantic, morpho-syntactic, syntactic, and discourse-pragmatic. The phonetic and phonological category includes features at the level of the speech sound such as pausing behaviour, and the time taken to produce components of speech including words, phonemes, and syllables.

Lexico-semantic features includes features at the word and content level, such as number of nouns, verbs, adjectives, and pronouns, obtained through techniques like part-of-speech tagging (Jarrold et al., 2020). The morphosyntactic-syntactic category includes features relating to inflectional morphology, while the syntactic category is features purely related to syntax, such as the number of words per clause, utterance, and sentence. Finally the discourse and pragmatic category was comprised of features that contribute to the continuation of conversation, such as cohesion, coherence, and correct use of conjunctions. (Boschi et al., 2017). An additional category for error typing was added, as recent research has demonstated the importance of these features in FTD (Bruffaerts et al., 2020; Catricalà et al., 2015). This additional category involved any features relating to the number or rate of a type of any errors.

44 of these features of speech and language were studied in more than one language and we focused on these features (see Table 1 for definitions). Within these, the most widely studied features were speech rate and articulation rate. The 18 features with the same main finding in more than one language are shown in Table 2. Six of the features were found to have the same main finding in both *more than one language* and in *more than one clinical variant of FTD*. For these features, the quantitative values of the papers were extracted and plotted for interlinguistic comparison in Figure 4. Quantitative value comparisons aim to identify potential variances in baseline measures among healthy controls across distinct languages, which may be important to determine differences in cut-off points for what should be considered normal values of different features within each language.

**Table 1.**
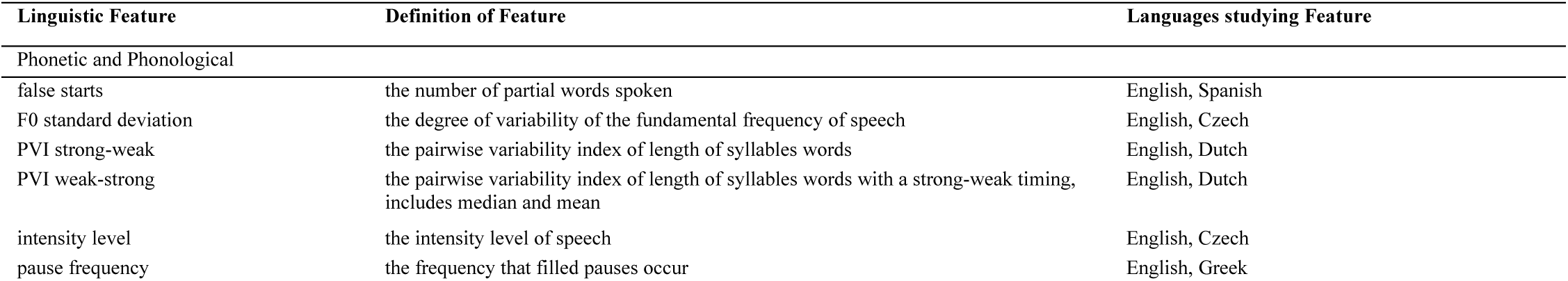
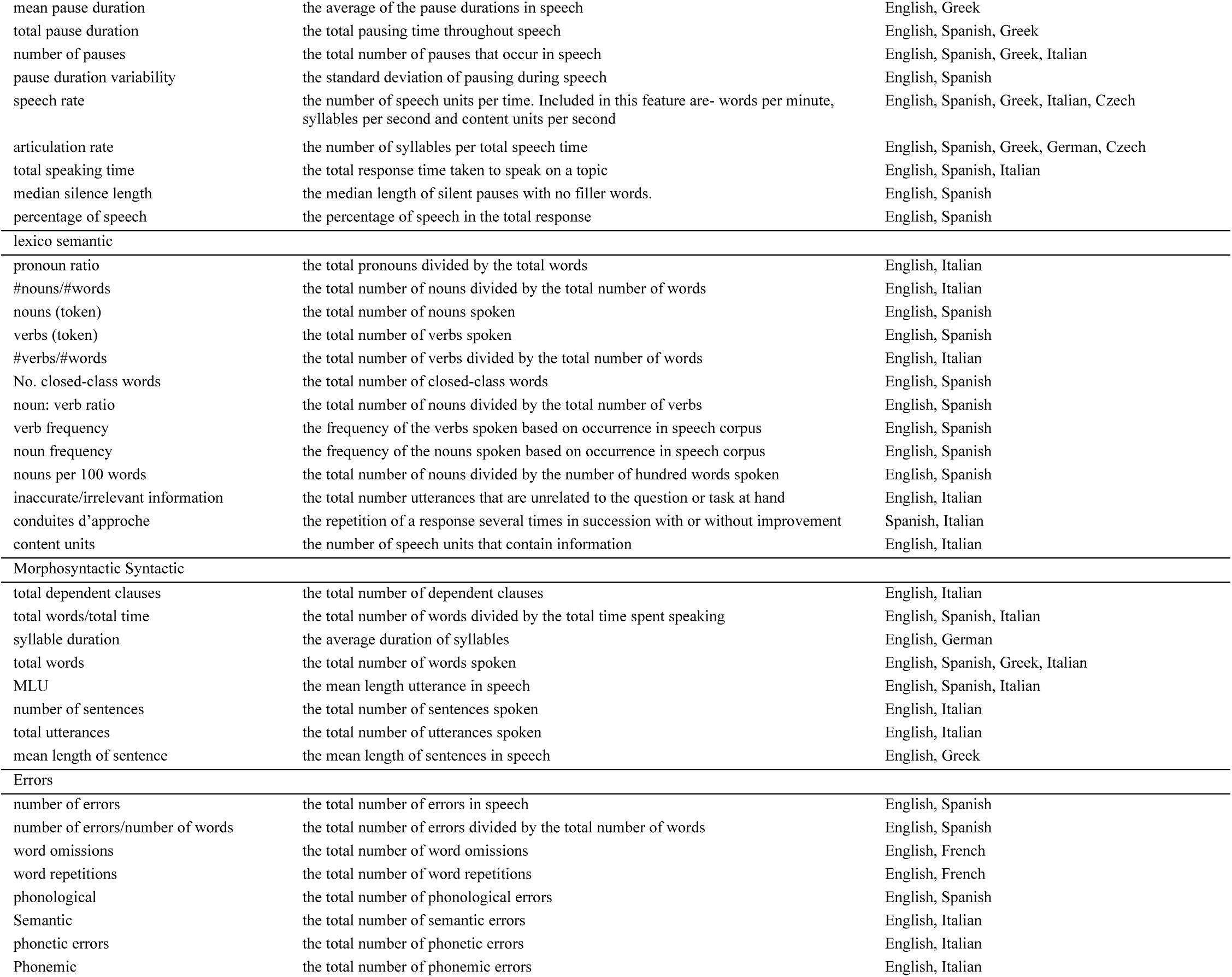
The definitions of the 44 linguistic and psychoacoustic features studied in more than one language in the FTD spectrum. References to the papers can be found in the supplementary materials.

**Table 2.**
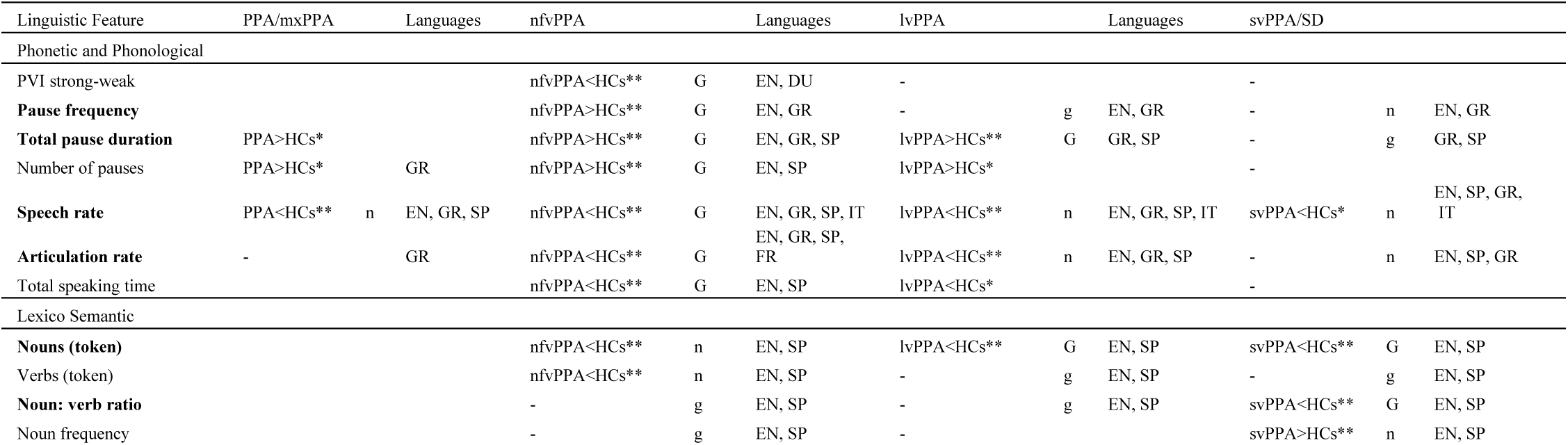
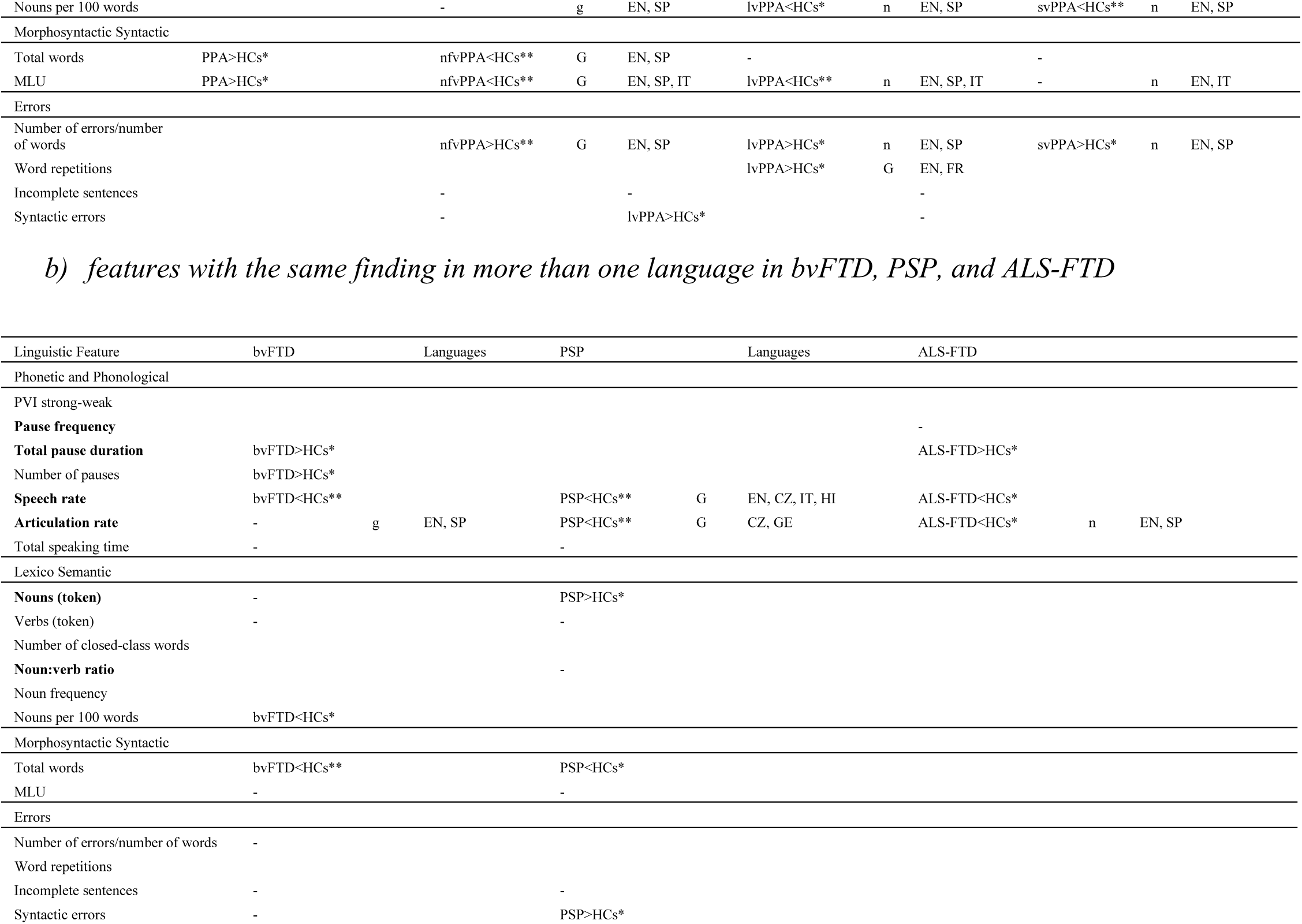
a) features with the same finding in more than one language in PPA.

*Note*: - attested in at least one paper and insignificant in more than half of papers; *attested and significant in one paper and significant in half or more of papers; **attested and significant in two or more papers and significant in half or more of the papers, g: same insignificant result in more than one language, G: same significant result in more than one language, n: different result in more than one language. The features in **bold** are generalisable in more than one variant.

**Figure 4.**
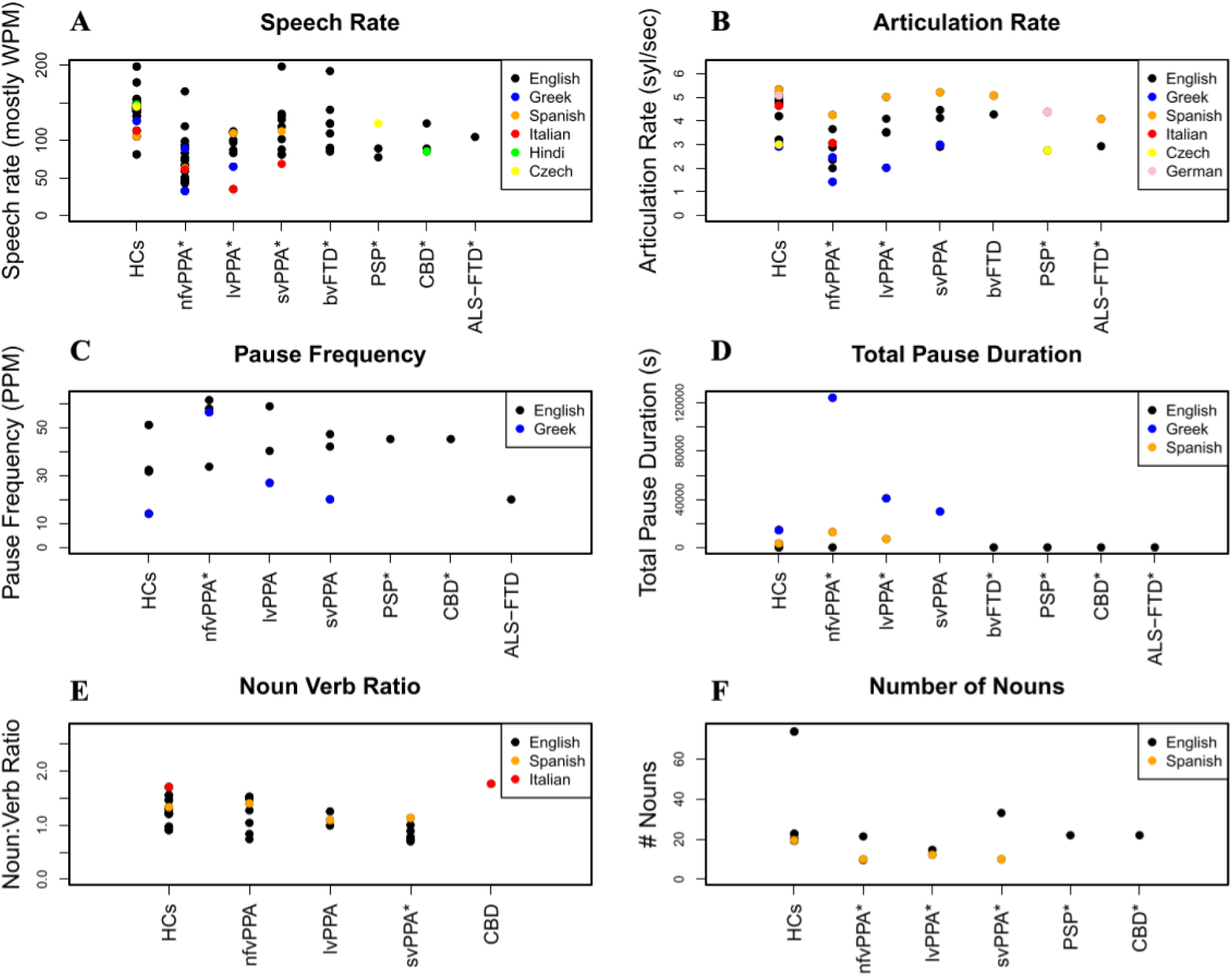
The quantitative values of each paper studying the 6 most generalizable features: speech rate, articulation rate, pause frequency, total pause duration, noun: verb ratio, and number of nouns.

*Note:* WPM = words per minute, PPM = pauses per minute. Different languages are represented by different colors. * Shows which groups were significantly different to controls (in at least half of papers). Certain papers used different units of measurement, for instance, some papers used pauses per second (PPS) while others used pauses per minute (PPM) (Figure 4C). In these instances, the PPS values were multiplied by 60 to compare them to the PPM values. Some features were not possible to transform, for instance, some papers investigated the number of nouns per number of total words whereas some investigated the number of nouns per total time. In this instance the values could not be transformed so only the number of nouns per time was included in the plot. For speech rate, the papers investigating syllables per second were combined with papers investigating words per minute (after being multiplied by 60). This does not appear to have affected the results, as the highest values for speech rate do not measure in syllables per second, but in words per minute. For articulation rate the papers using syllables per minute were divided by 60 to compare to the papers using syllables per second.

As is visible in Figure 4A, the **speech rate** was significantly reduced compared to controls in more than half the papers for all patient groups except ALS-FTD. However, the Greek patient populations with mxPPA, lvPPA, and svPPA had no significant reduction in speech rate (Karpathiou & Kambanaros, 2022; Potagas et al., 2022). Some English patient populations with lvPPA, svPPA, and bvFTD also had no significant differences in speech rate (Ash et al., 2013; Bird et al., 2000; Mack et al., 2015; Marcotte et al., 2017; Pressman et al., 2019; Rohrer & Warren, 2010; Sajjadi et al., 2012; Thompson, 2012; Wilson et al., 2010). Aside from Greek, all other non-English languages (Spanish, Italian, Hindi, and Czech) studying the FTD spectrum had a significant reduction in speech rate (Catricala et al., 2019; Daoudi et al., 2022; Matias-Guiu et al., 2022; Sachin et al., 2008; Silveri et al., 2012). The values of healthy controls do not visibly differ based on language. However, considerable variability within the English-speaking controls was observed.

Figure 4B shows that the **articulation rate** was reduced in more than half of papers investigating nfvPPA, lvPPA, and PSP. As is visible in Figure 4B, there was an overall trend of Greek patients and controls having the highest values for articulation rate and Spanish patient and control samples having the lowest values (Baque et al., 2022; Karpathiou & Kambanaros, 2022; Potagas et al., 2022). However, the lvPPA Spanish and Greek patient samples did not have a significant difference compared to healthy controls while the English patient samples were significantly reduced (Cho et al., 2022; Cordella et al., 2019; Cordella et al., 2017). PSP patients had a significantly reduced articulation rate in both Czech and German patient samples (Rusz et al., 2015; Skrabal et al., 2020; Staiger et al., 2017). ALS-FTD also had a significantly reduced articulation rate for English patients (Yunusova et al., 2016), however there was no significant difference for the Spanish patient sample (Baque et al., 2022). Finally, the bvFTD patient samples did not have a significant difference in articulation rate compared to controls for both the English and Spanish patient groups (Baque et al., 2022; Yunusova et al., 2016). The values of healthy controls do not visibly differ based on language.

The **pause frequency** was increased in nfvPPA in English-speaking patient samples as well as Greek (Cordella, 2017; Nevler et al., 2019; Parjane et al., 2021; Potagas et al., 2022). For lvPPA, svPPA, and ALS-FTD patient groups, the difference was insignificant in most papers in both Greek and English-speaking patient samples (Cordella et al., 2017; Potagas et al., 2022; Yunusova et al., 2016). For PSP and CBS, the pause frequency was also increased in one English patient sample (Parjane et al., 2021). The **total pause duration** was significantly increased in nfvPPA, lvPPA, bvFTD, PSP, CBS, and ALS-FTD in all English, Spanish, and Greek-speaking patient samples (Matias-Guiu et al., 2020; Parjane et al., 2021, 2021; Potagas et al., 2022; Yunusova et al., 2016). In one Greek svPPA sample, however, there was no significant difference in pause duration (Potagas et al., 2022). The values of healthy controls do not appear to differ based on language for total pause duration, however the Greek values were lower than the English for the pause frequency.

The **noun: verb ratio** was significantly reduced in most English patient groups with svPPA (Fraser et al., 2014; Garrard & Forsyth, 2010; Mack et al., 2015; Thompson, 2012). However, in one Spanish patient sample there was no significant difference in this ratio (Matias-Guiu et al., 2022). For the English, Spanish, and Italian patients with nfvPPA, lvPPA, and PSP there were no significant differences in the noun: verb ratio (Catricala et al., 2019; Fraser et al., 2014; Graham et al., 2004; Knibb et al., 2009; Mack et al., 2015; Marcotte et al., 2017; Matias-Guiu et al., 2022; Thompson et al., 1997; Thompson C.K., 2012). The **total number of nouns** was significantly decreased in nfvPPA, lvPPA, and svPPA, in both Spanish (Matias-Guiu et al., 2022) and English-speaking patients (Ash et al., 2009; Cho et al., 2022; Cupit et al., 2017; Fraser et al., 2014; Graham et al., 2004; Mack et al., 2015). In contrast, the total number of nouns was significantly increased in PSP and CBS in one English patient sample (Parjane et al., 2021). There was no significant difference in the number of nouns produced in bvFTD in one English-speaking patient sample (Ash et al., 2009). The values of healthy controls do not appear to differ based on language for the noun: verb ratio or the total number of nouns. Some variability was observed for noun: verb ratio in the English-speaking control group (the highest having a ratio of 1.55 (Fraser et al., 2014) and the lowest a ratio of 0.9 (Garrard & Forsyth, 2010)). the Italian-speaking control group had the highest noun: verb ratio.

Further information about the speech and language alterations found in FTD can be found in supplementary materials, which includes the sample sizes, languages, speech and language features studied, and relevant findings of all included papers.

## 4. DISCUSSION

This review shows that the most generalizable speech and language features of FTD across languages are speech rate, articulation rate, pause frequency, total pause duration, noun: verb ratio, and total number of nouns. Four out of the six features are in the phonetic and phonological category and the remaining two are in the lexico-semantic category. Phonetic and phonological features were useful in the detection of nfvPPA, and PSP. In contrast, lexico-semantic features were more able to detect svPPA. Our results suggest that connected speech analyses on relatively short samples can be used across multiple diverse languages to detect emerging neurodegenerative diseases in the FTD-spectrum. Our findings also promote the use of not just one single feature but advocate for the creation of an individualized speech and language profile for each phenotype combining both phonological and lexico-semantic features.

With regards to the phonetic and phonological features, one aspect that should be considered is the fact that nfvPPA is the most widely studied of the variants, both in number of papers and number of languages. nfvPPA tends to have more alterations at the speech sound level, in the phonetic and phonological category, which is reflected in the fact that such features were found to be generalizable. While phonetic and phonological features offer the most potential as interlinguistic markers of nfvPPA, inter-linguistic differences have been described in patients with nfvPPA. For instance, when comparing Italian nfvPPA patients to English nfvPPA patients, Canu et. al., 2020, found significant differences in the number of motor speech errors. In addition, work in Chinese patients with nfvPPA shows an increase in compound word production and in radical dysgraphia (substitution, transposition, omission, or addition of graphical units) relative to English patients (Tee et al., 2022). Despite differences, the findings of this review show generalizable results across language groups for phonetic and phonological features. The literature suggests that phonetic and phonological features may be broadly applicable in the Indo-European languages, in PSP as well as nfvPPA, with Greek and Hindi as Indo-Iranian languages (Potagas et al., 2022; Sachin et al., 2008), Czech as a Slavic language (Daoudi et al., 2022; Rusz et al., 2015; Skrabal et al., 2020), Italian, French, and Spanish as Romance languages (Baque et al., 2022; Bouvier et al., 2021; Silveri et al., 2014), and English and German as Germanic languages (Hohlbaum et al., 2018; Parjane et al., 2021).

The lexicosemantic features of speech and language were, as expected, the most studied and generalizable features for svPPA, though they are only studied in Indo-European languages. However, lexicosemantic features also offer the potential to diagnose lvPPA. Due to its underlying pathology, lvPPA is often considered an atypical variant of Alzheimer’s Disease (AD), rather than a variant of FTD. This could have constituted a reason to exclude such cases in the present review. However, as lvPPA patients are an integral clinical phenotype within the primary progressive aphasias, and it is often a necessary challenge to differentiate nfvPPA and lvPPA at an early stage, we chose to include these cases. Moreover, findings of speech and language differences in lvPPA may inform the study of typical AD patients. For instance, the number of nouns is reduced in connected speech in both AD and lvPPA (Fraser et al., 2016; Matias-Guiu et al., 2020). Pronoun use is also increased in both lvPPA and AD (Boschi et al., 2017; Lavoie et al., 2021; Slegers et al., 2018; Wilson et al., 2010). In addition, impaired naming abilities are typically found in both disorders (Brandt et al., 2010; Evrard, 2002; Jebahi et al., 2023).

Morphosyntactic and syntactic features tend to differ across languages so it was expected that these features would be less generalizable, though they are associated with nfvPPA which was most frequently studied. Discourse and pragmatic features were not widely studied in the reviewed papers, and definitions of cohesiveness and coherence varied greatly. However, with consistent measures, this category may still contain generalizable features of the FTD spectrum.

Defining the variants of PPA poses a challenge in the clinic, as the optimal diagnostic tools and definitions of certain speech and language abnormalities are still subject to debate. It is also likely that the criteria for PPA derived in English-speaking patients do not perfectly map onto those of non-English-speaking PPA patients (Tee et al., 2022). For instance, word repetition tasks with polysyllabic words with consonant clusters often used in English to detect PPAOS, are not applicable for Chinese patients, as Cantonese is generally monosyllabic (Tee et al., 2022). Even within English, there is some contention regarding aspects such as motor speech, which has variable definitions depending on the authors and clinicians (Duffy et al., 2014; Grossman, 2018).

While the Indo-European languages are relatively well represented in the papers included in this review, data is lacking entirely for some major language groups. At the time of writing there are no known papers studying connected speech in FTD for the Uralic, Altaic, Caucasian, Sino-Tibetan, Tai, Austronesian, Niger-Congo, or Afro-Asiatic language families. The absence of diverse language representation in FTD research hinders tailored characterization and sensitive assessment methods for non-English speaking patients. This gap could result in delayed or misdiagnosis, and ultimately perpetuate healthcare disparities. However, some research is being carried out investigating language in FTD for these language groups, for instance the Genetic FTD Initiative (GENFI) consortium is starting to investigate Finnish (a Uralic language). Further research is necessary to determine whether the results from Indo-European languages apply to these other major language groups.

Another consideration when investigating speech and language markers is the difference in quantitative values across different languages, as well as the variation seen within languages. It is difficult to discern which differences are due to linguistic differences and which are due to differences in methods and metrics. Differences in task use likely contributes to the observed variation within English controls. In addition, FTD is a very heterogenous group, and disease stage plays a great role. Different studies often include patients with different disease severities, which may also be a contributor to the observed variation within English patients. Standardization using z-scores based on norms for each language would make comparison across languages feasible and allow us to apply findings to less studied language groups. On the other hand, simply translating speech and language tests into another language result is not always possible: for every language, linguistically and culturally equivalent tasks are necessary and should be developed to elicit valid responses (Fyndanis et al., 2017).

In alignment with the findings of García et al., 2023 on speech and language research in neurodegenerative diseases, we reiterate the need for cross-linguistic behavioural research in the FTD spectrum, especially considering English is not the optimal baseline for all languages. Collection of data in a standardized and transparent way is vital for future comparison across languages and disorders, and to determine the correct baseline values for different disorders. This requires the adaptation of tasks for the study of connected speech to other languages. We show a clear need for further investigation of speech and language markers of the FTD spectrum in more non-English languages, especially non-Indo-European languages.

## 5. CONCLUSION

The findings of this systematic review show that while interlinguistic differences in FTD patients exist, there may indeed be features of speech and language that are generalizable across several languages. Namely phonological and lexico-semantic features, offer the potential for future implementation as interlinguistic markers of FTD. Further study of these variables in different languages and across the FTD spectrum will determine the applicability of these markers in the clinic.

## Data Availability

All data produced in the present work are contained in the manuscript

## REFERENCES

Ash, S., Evans, E., O’Shea, J., Powers, J., Boller, A., Weinberg, D., Haley, J., McMillan, C., Irwin, D. J., Rascovsky, K., & Grossman, M. (2013). Differentiating primary progressive aphasias in a brief sample of connected speech. Neurology, 81(4), 329–336. Scopus. 10.1212/WNL.0b013e31829c5d0e

Ash, S., Moore, P., Antani, S., McCawley, G., Work, M., & Grossman, M. (2006). Trying to tell a tale: Discourse impairments in progressive aphasia and frontotemporal dementia. Neurology, 66(9), 1405–1413. 10.1212/01.wnl.0000210435.72614.38

Ash, S., Moore, P., Vesely, L., Gunawardena, D., McMillan, C., Anderson, C., Avants, B., & Grossman, M. (2009). Non-Fluent Speech in Frontotemporal Lobar Degeneration. Journal of Neurolinguistics, 22(4), 370–383. 10.1016/j.jneuroling.2008.12.001

Baque, L., Machuca, M. J., & Santos-Santos, M. A. (2022). Preliminary study of the temporal variables of continuous speech in patients with neurodegenerative syndromes of the frontotemporal lobar degeneration spectrum. Revista de Neurologia, 74(2), 37–47. 10.33588/rn.7402.2021197

Bird, H., Lambon Ralph, M. A., Patterson, K., & Hodges, J. R. (2000). The Rise and Fall of Frequency and Imageability: Noun and Verb Production in Semantic Dementia. Brain and Language, 73(1), 17–49. 10.1006/brln.2000.2293

Boeve, B. F., Boxer, A. L., Kumfor, F., Pijnenburg, Y., & Rohrer, J. D. (2022). Advances and controversies in frontotemporal dementia: Diagnosis, biomarkers, and therapeutic considerations. The Lancet Neurology, 21(3), 258–272. 10.1016/S1474-4422(21)00341-0

Boschi, V., Catricalà, E., Consonni, M., Chesi, C., Moro, A., & Cappa, S. F. (2017). Connected Speech in Neurodegenerative Language Disorders: A Review. Frontiers in Psychology, 8, 269. 10.3389/fpsyg.2017.00269

Bouvier, L., Monetta, L., Laforce, R., Jr., Vitali, P., Bocti, C., & Martel-Sauvageau, V. (2021). Progressive apraxia of speech in Quebec French speakers: A case series. International Journal of Language & Communication Disorders, 56(3), 528–548. 10.1111/1460-6984.12606

Brandt, J., Bakker, A., & Maroof, D. A. (2010). Auditory confrontation naming in alzheimer’s disease. The Clinical Neuropsychologist, 24(8), 1326–1338. 10.1080/13854046.2010.518977

Bruffaerts, R., Schaeverbeke, J., De Weer, A.-S., Nelissen, N., Dries, E., Van Bouwel, K., Sieben, A., Bergmans, B., Swinnen, C., Pijnenburg, Y., Sunaert, S., Vandenbulcke, M., & Vandenberghe, R. (2020). Multivariate analysis reveals anatomical correlates of naming errors in primary progressive aphasia. Neurobiology of Aging, 88, 71–82. 10.1016/j.neurobiolaging.2019.12.016

Bruffaerts, R., Schaeverbeke, J., Radwan, A., Grube, M., Gabel, S., De Weer, A.-S., Dries, E., Van Bouwel, K., Griffiths, T. D., Sunaert, S., & Vandenberghe, R. (2022). Left Frontal White Matter Links to Rhythm Processing Relevant to Speech Production in Apraxia of Speech. Neurobiology of Language, 3(4), 515–537. 10.1162/nol_a_00075

Canu, E., Agosta, F., Battistella, G., Spinelli, E. G., DeLeon, J., Welch, A. E., … & Gorno-Tempini, M. L. (2020). Speech production differences in English and Italian speakers with nonfluent variant PPA. Neurology, 94(10), e1062–e1072.

Catricala, E., Boschi, V., Cuoco, S., Galiano, F., Picillo, M., Gobbi, E., Miozzo, A., Chesi, C., Esposito, V., Santangelo, G., Pellecchia, M. T., Borsa, V. M., Barone, P., Garrard, P., Iannaccone, S., & Cappa, S. F. (2019). The language profile of progressive supranuclear palsy. Cortex, 115, 294–308. 10.1016/j.cortex.2019.02.013

Catricalà, E., Della Rosa, P. A., Plebani, V., Perani, D., Garrard, P., & Cappa, S. F. (2015). Semantic feature degradation and naming performance. Evidence from neurodegenerative disorders. Brain and Language, 147, 58–65. 10.1016/j.bandl.2015.05.007

Cho, S., Cousins, K. A. Q., Shellikeri, S., Ash, S., Irwin, D. J., Liberman, M. Y., Grossman, M., & Nevler, N. (2022). Lexical and Acoustic Speech Features Relating to Alzheimer Disease Pathology. Neurology, 99(4), E313–E322. 10.1212/WNL.0000000000200581

Cordella, C., Dickerson, B. C., Quimby, M., Yunusova, Y., & Green, J. R. (2017). Slowed articulation rate is a sensitive diagnostic marker for identifying non-fluent primary progressive aphasia. Aphasiology, 31(2), 241–260.

Cordella, C., Quimby, M., Touroutoglou, A., Brickhouse, M., Dickerson, B. C., & Green, J. R. (2019). Quantification of motor speech impairment and its anatomic basis in primary progressive aphasia. Neurology, 92(17), e1992–e2004. 10.1212/WNL.0000000000007367

Cupit, J., Leonard, C., Graham, N. L., Lima, B. S., Tang-Wai, D., Black, S. E., & Rochon, E. (2017). Analysing syntactic productions in semantic variant PPA and non-fluent variant PPA: how different are they? Aphasiology, 31(3), 282–307. 10.1080/02687038.2016.1180661

Daoudi, K., Das, B., Tykalova, T., Klempir, J., & Rusz, J. (2022). Speech acoustic indices for differential diagnosis between Parkinson’s disease, multiple system atrophy and progressive supranuclear palsy. Npj Parkinson’s Disease, 8(1). Scopus. 10.1038/s41531-022-00389-6

Duffy, J. R., Strand, E. A., & Josephs, K. A. (2014). Motor Speech Disorders Associated with Primary Progressive Aphasia. Aphasiology, 28(8–9), 1004–1017. 10.1080/02687038.2013.869307

Evrard, M. (2002). Ageing and lexical access to common and proper names in picture naming. Brain and Language, 81(1–3), 174–179. 10.1006/brln.2001.2515

Fraser, K. C., Meltzer, J. A., Graham, N. L., Leonard, C., Hirst, G., Black, S. E., & Rochon, E. (2014). Automated classification of primary progressive aphasia subtypes from narrative speech transcripts. Cortex, 55, 43–60. 10.1016/j.cortex.2012.12.006

Fraser, K. C., Meltzer, J. A., & Rudzicz, F. (2016). Linguistic Features Identify Alzheimer’s Disease in Narrative Speech. Journal of Alzheimers Disease, 49(2), 407–422. 10.3233/JAD-150520

Fyndanis, V., Lind, M., Varlokosta, S., Kambanaros, M., Soroli, E., Ceder, K., Grohmann, K. K., Rofes, A., Simonsen, H. G., Bjekić, J., Gavarró, A., Kuvač Kraljević, J., Martínez-Ferreiro, S., Munarriz, A., Pourquie, M., Vuksanović, J., Zakariás, L., & Howard, D. (2017). Cross-linguistic adaptations of The Comprehensive Aphasia Test: Challenges and solutions. Clinical Linguistics & Phonetics, 31(7–9), 697–710. 10.1080/02699206.2017.1310299

García, A. M., de Leon, J., Tee, B. L., Blasi, D. E., & Gorno-Tempini, M. L. (2023). Speech and language markers of neurodegeneration: A call for global equity. Brain, awad 253. 10.1093/brain/awad253

Garrard, P., & Forsyth, R. (2010). Abnormal discourse in semantic dementia: A data-driven approach. Neurocase, 16(6), 520–528. 10.1080/13554791003785901

Geraudie, A., Battista, P., García, A. M., Allen, I. E., Miller, Z. A., Gorno-Tempini, M. L., & Montembeault, M. (2021). Speech and language impairments in behavioral variant frontotemporal dementia: A systematic review. Neuroscience & Biobehavioral Reviews, 131, 1076–1095. 10.1016/j.neubiorev.2021.10.015

Goodglass, H., & Kaplan, E. (1972). The assessment of aphasia and related disorders. Lea & Febiger.

Gorno-Tempini, M. L., Hillis, A. E., Weintraub, S., Kertesz, A., Mendez, M., Cappa, S. F., Ogar, J. M., Rohrer, J. D., Black, S., Boeve, B. F., Manes, F., Dronkers, N. F., Vandenberghe, R., Rascovsky, K., Patterson, K., Miller, B. L., Knopman, D. S., Hodges, J. R., Mesulam, M. M., & Grossman, M. (2011). Classification of primary progressive aphasia and its variants. Neurology, 76(11), Article 11. 10.1212/WNL.0b013e31821103e6

Graham, N., Patterson, K., & Hodges, J. (2004). When more yields less: Speaking and writing deficits in Nonfluent progressive aphasia. Neurocase, 10(2), 141–155. 10.1080/13554790490497256

Grossman, M. (2018). Linguistic Aspects of Primary Progressive Aphasia. In M. Liberman & B. Partee (Eds.), Annual Review of Linguistics, *VOL 4* (Vol. 4, pp. 377–403). 10.1146/annurev-linguistics-011516-034253

Hohlbaum, K., Dressel, K., Lange, I., Wellner, B., Saez, L. E., Huber, W., Grande, M., Amunts, K., Grodzinsky, Y., & Heim, S. (2018). Sentence repetition deficits in the logopenic variant of PPA: linguistic analysis of longitudinal and cross-sectional data. Aphasiology, 32(12), 1445–1467. 10.1080/02687038.2017.1423271

Jebahi, F., Nickels, K. V., & Kielar, A. (2023). Predicting Confrontation Naming in the Logopenic Variant of Primary Progressive Aphasia. Aphasiology, 0(0), 1–32. 10.1080/02687038.2023.2221998

Karpathiou, N., & Kambanaros, M. (2022). Comparing Individuals With PPA to Individuals With AD: Cognitive and Linguistic Profiles. Frontiers in Communication, 7. Scopus. 10.3389/fcomm.2022.893471

Knibb, J. A., Woollams, A. M., Hodges, J. R., & Patterson, K. (2009). Making sense of progressive non-fluent aphasia: An analysis of conversational speech. Brain : A Journal of Neurology, 132(Pt 10), 2734–2746. 10.1093/brain/awp207

Koukoulioti, V., Stavrakaki, S., Konstantinopoulou, E., & Ioannidis, P. (2018). Lexical and Grammatical Factors in Sentence Production in Semantic Dementia: Insights From Greek. Journal of Speech, Language, and Hearing Research : JSLHR, 61(4), 870–886. 10.1044/2017_JSLHR-L-17-0024

Koukoulioti, V., Stavrakaki, S., Konstantinopoulou, E., & Ioannidis, P. (2020). Time reference, morphology and prototypicality: Tense production in stroke aphasia and semantic dementia in Greek. Clinical Linguistics & Phonetics, 34(9), 791–825. 10.1080/02699206.2019.1700308

Lavoie, M., Black, S. E., Tang-Wai, D. F., Graham, N. L., Stewart, S., Leonard, C., & Rochon, E. (2021). Description of connected speech across different elicitation tasks in the logopenic variant of primary progressive aphasia. International Journal of Language & Communication Disorders, 56(5), 1074–1085. 10.1111/1460-6984.12660

Mack, J. E., Chandler, S. D., Meltzer-Asscher, A., Rogalski, E., Weintraub, S., Mesulam, M.-M., & Thompson, C. K. (2015). What do pauses in narrative production reveal about the nature of word retrieval deficits in PPA? Neuropsychologia, 77, 211–222. 10.1016/j.neuropsychologia.2015.08.019

Macoir, J., Martel-Sauvageau, V., Bouvier, L., Laforce, R., & Monetta, L. (2021). Heterogeneity of repetition abilities in logopenic variant primary progressive aphasia. Dementia & Neuropsychologia, 15(3), 405–412. 10.1590/1980-57642021dn15-030014

Marcotte, K., Graham, N. L., Fraser, K. C., Meltzer, J. A., Tang-Wai, D. F., Chow, T. W., Freedman, M., Leonard, C., Black, S. E., & Rochon, E. (2017). White Matter Disruption and Connected Speech in Non-Fluent and Semantic Variants of Primary Progressive Aphasia. Dementia and Geriatric Cognitive Disorders Extra, 7(1), 52–73. 10.1159/000456710

Matias-Guiu, J. A., Suárez-Coalla, P., Pytel, V., Cabrera-Martín, M. N., Moreno-Ramos, T., Delgado-Alonso, C., Delgado-Álvarez, A., Matías-Guiu, J., & Cuetos, F. (2020). Reading prosody in the non-fluent and logopenic variants of primary progressive aphasia. Cortex, 132, 63–78. 10.1016/j.cortex.2020.08.013

Matias-Guiu, J. A., Suarez-Coalla, P., Yus, M., Pytel, V., Hernandez-Lorenzo, L., Delgado-Alonso, C., Delgado-Alvarez, A., Gomez-Ruiz, N., Polidura, C., Nieves Cabrera-Martin, M., Matias-Guiu, J., & Cuetos, F. (2022). Identification of the main components of spontaneous speech in primary progressive aphasia and their neural underpinnings using multimodal MRI and FDG-PET imaging. Cortex, 146, 141–160. 10.1016/j.cortex.2021.10.010

Moore, K. M., Nicholas, J., Grossman, M., McMillan, C. T., Irwin, D. J., Massimo, L., Van Deerlin, V. M., Warren, J. D., Fox, N. C., Rossor, M. N., Mead, S., Bocchetta, M., Boeve, B. F., Knopman, D. S., Graff-Radford, N. R., Forsberg, L. K., Rademakers, R., Wszolek, Z. K., van Swieten, J. C., … Geschwind, D. (2020). Age at symptom onset and death and disease duration in genetic frontotemporal dementia: An international retrospective cohort study. The Lancet Neurology, 19(2), Article 2. 10.1016/S1474-4422(19)30394-1

Nevler, N., Ash, S., Irwin, D. J., Liberman, M., & Grossman, M. (2019). Validated automatic speech biomarkers in primary progressive aphasia. Annals of Clinical and Translational Neurology, 6(1), 4–14. 10.1002/acn3.653

Ouzzani, M., Hammady, H., Fedorowicz, Z., & Elmagarmid, A. (2016). Rayyan—A web and mobile app for systematic reviews. Systematic Reviews, 5(1), 210. 10.1186/s13643-016-0384-4

Parjane, N., Cho, S., Ash, S., Cousins, K. A. Q., Shellikeri, S., Liberman, M., Shaw, L. M., Irwin, D. J., Grossman, M., & Nevler, N. (2021). Digital Speech Analysis in Progressive Supranuclear Palsy and Corticobasal Syndromes. Journal of Alzheimers Disease, 82(1), 33–45. 10.3233/JAD-201132

Peterson, K. A., Patterson, K., & Rowe, J. B. (2019). Language impairment in progressive supranuclear palsy and corticobasal syndrome. Journal of Neurology. 10.1007/s00415-019-09463-1

Potagas, C., Nikitopoulou, Z., Angelopoulou, G., Kasselimis, D., Laskaris, N., Kourtidou, E., … & Kapaki, E. (2022). Silent Pauses and Speech Indices as Biomarkers for Primary Progressive Aphasia. Medicina, 58(10), 1352.

Pressman, P. S., Ross, E. D., Cohen, K. B., Chen, K.-H., Miller, B. L., Hunter, L. E., Gorno-Tempini, M. L., & Levenso, R. W. (2019). Interpersonal prosodic correlation in frontotemporal dementia. Annals of Clinical and Translational Neurology, 6(7), 1352–1357. 10.1002/acn3.50816

Rohrer, J. D., Nicholas, J. M., Cash, D. M., van Swieten, J., Dopper, E., Jiskoot, L., van Minkelen, R., Rombouts, S. A., Cardoso, M. J., Clegg, S., Espak, M., Mead, S., Thomas, D. L., De Vita, E., Masellis, M., Black, S. E., Freedman, M., Keren, R., MacIntosh, B. J., … Binetti, G. (2015). Presymptomatic cognitive and neuroanatomical changes in genetic frontotemporal dementia in the Genetic Frontotemporal dementia Initiative (GENFI) study: A cross-sectional analysis. The Lancet. Neurology, 14(3), Article 3. 10.1016/S1474-4422(14)70324-2

Rohrer, J. D., & Warren, J. D. (2010). Phenomenology and anatomy of abnormal behaviours in primary progressive aphasia. Journal of the Neurological Sciences, 293(1–2), 35–38. 10.1016/j.jns.2010.03.012

Rusina, R., Vandenberghe, R., & Bruffaerts, R. (2021). Cognitive and Behavioral Manifestations in ALS: Beyond Motor System Involvement. Diagnostics, 11(4), 624. 10.3390/diagnostics11040624

Rusz, J., Bonnet, C., Klempíř, J., Tykalová, T., Baborová, E., Novotný, M., Rulseh, A., & Růžička, E. (2015). Speech disorders reflect differing pathophysiology in Parkinson’s disease, progressive supranuclear palsy and multiple system atrophy. Journal of Neurology, 262(4), 992–1001. Scopus. 10.1007/s00415-015-7671-1

Sachin, S., Shukla, G., Goyal, V., Singh, S., Aggarwal, V., Gureshkumar, & Behari, M. (2008). Clinical speech impairment in Parkinson’s disease, progressive supranuclear palsy, and multiple system atrophy. Neurology India, 56(2), 122–126. Scopus. 10.4103/0028-3886.41987

Sajjadi, S. A., Patterson, K., Tomek, M., & Nestor, P. J. (2012). Abnormalities of connected speech in the non-semantic variants of primary progressive aphasia. Aphasiology, 26(10), 1219–1237. Scopus. 10.1080/02687038.2012.710318

Samra, K., MacDougall, A. M., Bouzigues, A., Bocchetta, M., Cash, D. M., Greaves, C. V., Convery, R. S., van Swieten, J. C., Seelaar, H., Jiskoot, L., Moreno, F., Sanchez-Valle, R., Laforce, R., Graff, C., Masellis, M., Tartaglia, M. C., Rowe, J. B., Borroni, B., Finger, E., … Russell, L. L. (2023). Language impairment in the genetic forms of behavioural variant frontotemporal dementia. Journal of Neurology, 270(4), 1976–1988. 10.1007/s00415-022-11512-1

Silveri, M. C., Ciccarelli, N., Baldonero, E., Piano, C., Zinno, M., Soleti, F., Bentivoglio, A. R., Albanese, A., & Daniele, A. (2012). Effects of stimulation of the subthalamic nucleus on naming and reading nouns and verbs in Parkinson’s disease. Neuropsychologia, 50(8), 1980–1989. 10.1016/j.neuropsychologia.2012.04.023

Silveri, M. C., Pravata, E., Brita, A. C., Improta, E., Ciccarelli, N., Rossi, P., & Colosimo, C. (2014). Primary progressive aphasia: Linguistic patterns and clinical variants. Brain and Language, 135, 57–65. 10.1016/j.bandl.2014.05.004

Skrabal, D., Tykalova, T., Klempir, J., Ruzicka, E., & Rusz, J. (2020). Dysarthria enhancement mechanism under external clear speech instruction in Parkinson’s disease, progressive supranuclear palsy and multiple system atrophy. Journal of Neural Transmission, 127, 905–914.

Slegers, A., Filiou, R.-P., Montembeault, M., & Brambati, S. M. (2018). Connected Speech Features from Picture Description in Alzheimer’s Disease: A Systematic Review. Journal of Alzheimer’s Disease: JAD, 65(2), 519–542. 10.3233/JAD-170881

Staiger, A., Schölderle, T., Brendel, B., & Ziegler, W. (2017). Dissociating oral motor capabilities: Evidence from patients with movement disorders. Neuropsychologia, 95, 40–53.

Suh, M. K., Kim, E.-J., Lee, B. H., Seo, S. W., Chin, J., Kang, S. J., & Na, D. L. (2010). Hanja (Ideogram) alexia and agraphia in patients with semantic dementia. Neurocase, 16(2), 146–156. 10.1080/13554790903339629

Tee, B. L., Lorinda Kwan-Chen, L. Y., Chen, T. F., Yan, C. T., Tsoh, J., Lung-Tat Chan, A., … & Gorno-Tempini, M. L. (2022). Dysgraphia phenotypes in native Chinese speakers with primary progressive aphasia. Neurology, 98(22), e2245–e2257.

Thompson, C. K., Ballard, K. J., Tait, M. E., Weintraub, S., & Mesulam, M. (1997). Patterns of language decline in non-fluent primary progressive aphasia. Aphasiology, 11(4–5), 297–321. 10.1080/02687039708248473

Thompson, C. K., Lukic, S., King, M. C., Mesulam, M. M., & Weintraub, S. (2016). Verb and noun deficits in stroke-induced and primary progressive aphasia: The Northwestern Naming Battery. In The Science of Aphasia Rehabilitation (pp. 18-41). Routledge.

Wilson, S. M., Henry, M. L., Besbris, M., Ogar, J. M., Dronkers, N. F., Jarrold, W., Miller, B. L., & Gorno-Tempini, M. L. (2010). Connected speech production in three variants of primary progressive aphasia. Brain, 133(7), 2069–2088. 10.1093/brain/awq129

Yunusova, Y., Graham, N. L., Shellikeri, S., Phuong, K., Kulkarni, M., Rochon, E., Tang-Wai, D. F., Chow, T. W., Black, S. E., Zinman, L. H., & Green, J. R. (2016). Profiling Speech and Pausing in Amyotrophic Lateral Sclerosis (ALS) and Frontotemporal Dementia (FTD). Plos One, 11(1). 10.1371/journal.pone.0147573

