## Supplemental Table 1 for "A Systematic Review of the Quantitative markers of speech and language of the Frontotemporal Degeneration Spectrum and their potential for cross-linguistic implementation"

**Supplementary materials Table 1.** HCs : Healthy controls, PPA : Primary progressive aphasia, nfvPPA : non-fluent variant PPA, svPPA : semantic variant PPA, lvPPA : logopenic variant PPA, FTD : Frontotemporal dementia, bvFTD : Behavioural variant FTD, PSP : Progressive supranuclear palsy, CBS : Corticobasal degeneration, ALS-FTD : Amyotrophic lateral sclerosis with FTD.

| Year | First Author | Language | Participants | Methods | Speech and Language features | Results (significant only) |
| --- | --- | --- | --- | --- | --- | --- |
| 2022 | Daoudi | Czech | 150 HCs, 20 PSP | reading task, 90s monologue on chosen theme. | fundamental frequency variability (F0 SD), relative loudness of respiration (RLR), rate of speech respiration (RSR), pause intervals per respiration (PIR), gaping in-between voiced intervals (GVI), duration of stop consonants (DUS), resonance frequency attenuation (RFA), entropy of speech timing (EST), rate of speech timing (RST) | RLRm - increased in PSP, RSRm/t - increased in PSP, PIRm - decreased in PSP, PIRt - decreased in PSP, GVIm/t - decreased in PSP, DUSm - increased in PSP, DUSt (increased in PSP), ESTm - decreased in PSP, ESTt - decreased in PSP, RSTm - decreased in PSP, RSTt - decreased in PSP, ASTt - decreased in PSP, stdF0t - increased in PSP, NSRt - decreased in PSP. (t: text, m: monologue). |
| 2022 | Potagas | Greek | 17 HCs, 6 nfvPPA, 6 svPPA, 8 lvPPA | Picture description - cookie theft, personal narration - describe of course of disease | Pause frequency, pause total duration, speech rate, articulation rate | pause frequency (picture description) - increased in nfvPPA, lvPPA, and svPPA, pause frequency (personal narration) - increased in nfvPPA, lvPPA, pause total duration (picture description) - increased in nfvPPA, lvPPA, pause total duration (personal narration) - increased in nfvPPA, speech rate (picture description) - decreased in nfvPPA, lvPPA, speech rate (personal narration) - decreased in nfvPPA, lvPPA, articulation rate (picture description) - decreased in nfvPPA, lvPPA, articulation rate (picture description) - decreased in nfvPPA |
| 2022 | Garcia | English | 10 HCs, 22 nfvPPA, 15 svPPA | reading task - grandfather passage | Articulation rate, syllable duration, pause duration, syllable duration variability, pause duration variability | articulation rate – decreased in nfvPPA and svPPA, syllable duration – increased in nfvPPA and svPPA, pause duration – increased in nfvPPA and svPPA, syllable duration variability – increased in nfvPPA and svPPA, pause duration variability - increased in nfvPPA and svPPA |
| 2021 | Macoir | Quebec French | 12 HCs, 4 lvPPA | repetition subtests of the batterie d’évaluation cognitive du langage (battery of cognitive assessment of language), test français de répétition de phrases (French sentence repetition test) | repetition accuracy (error typing) | word omissions - increased in lvPPA, word additions - increased in lvPPA, and word repetitions - increased in lvPPA |
| 2021 | Parjane | English | 41 HCs, 25 nfvPPA, 87 PSP-CBD | Picture description - cookie theft | mean speech segment duration, mean pause segment duration, and pause rate, fundamental frequency (f0), Speech rate, dependent clauses per 100 utterances (DC) and percentage of utterances that are well-formed sentences (WFS). | nfvPPA and PSPS-CBS speech produced shorter speech segments, longer pauses segment durations, higher pause rates, reduced fundamental frequency (f0) pitch ranges, and slower speech rate compared to HCs. WPM was lower in both patient groups, verb production was lower in nfvPPA and insignificant in PSP-CBS, noun production lower in PSP-CBS and insignificant in nfvPPA. |
| 2020 | Skrabal | Czech | 17 HCs, 17 PSP | reading task - normally and in clear speech condition | intensity level, intensity variability, fundamental frequency variability (F0 SD), vowel space area (VSA), and articulation rate | F0 SD, VSA, and articulation rate - reduced in PSP. |
| 2019 | Pressman | English | 170 HCs, 29 bvFTD, 14 rtFTD, 14 nfvPPA, 19 svPPA | conversation | speech rate (syllables/second), variation of fundamental frequency, and standard deviation of intensity | speech rate (syllables/second) - reduced in bvFTD, nfvPPA. |
| 2019 | Nevler | English | 31 HCs, 15 nfvPPA, 21 svPPA, 23 lvPPA | picture description - cookie theft | fundamental frequency, speech and pause durations. | fundamental frequency - reduced in nfvPPA, pause rate - reduced in nfvPPA, svPPA, and lvPPA, mean speech duration - reduced in nfvPPA, svPPA, and lvPPA |
| 2018 | Hohlbaum | German | 7 HCs, 8 lvPPA | repetition task with: (a) simple S-V-O sentences, (b) sentences with a temporal adverb, (c) questions | sentence repetition errors | repetition errors - increased in lvPPA |
| 2017 | Vogel | English | 24 bvFTD, 24 HCs | 40s monologue, reading task - grandfather passage, days of the week | Mean silence length, variability of silence length, proportion of silence time, speech rate (syl/s) | Mean silence length - increased for bvFTD for "days of the week", unprepared monologue, variability of silence length - increased for bvFTD for unprepared monologue, proportion of silence time - increased for bvFTD for reading task and unprepared monologue, speech rate (syl/s) - decreased for reading passage |
| 2017 | Nevler | English | 17 HCs, 32 bvFTD | picture description - cookie theft | fundamental frequency - extracted pitch range, mean speech duration, mean pause duration, speech rate (WPM) | f0 range - decreased in bvFTD, speech duration - reduced in bvFTD, pause duration - increased in bvFTD, speech rate - reduced in bvFTD |
| 2017 | Cupit | English | 23 HCs, 18 nfvPPA, 13 svPPA | constrained picture description task, eliciting active, passive, dative, and dative–passive sentences | sentence production - noun, verb and grammar production | noun accuracy - reduced in nfvPPA and svPPA, difficulty with the passive and dative passive structures vs active and dative sentence structures - increased in nfvPPA and svPPA |
| 2017 | Marcotte | English | 18 HCs, 13 nfvPPA, 12 svPPA | topic-directed interviews - about (1) family, (2) health, (3) daily activities, (4) where they were born and raised, and (5) jobs or work | speech rate, number of sentences, mean length of sentence, mean word length, pronoun ratio, #nouns/#words, #verbs/#words, proportion verbs, noun: verb ratio, noun and verb frequency, imageability, adverbs, brunets index, words used once, type token ratio, mean length of clause, clauses, complex nominals per clause, T-units, mean length of T-unit, insignificant in svPPA, complex nominals per T-unit, total Yngve depth, max and mean Yngve depth | speech rate - reduced in nfvPPA, number of sentences - increased in nfvPPA, mean length of sentence - reduced in nfvPPA, mean word length - reduced in nfvPPA, pronoun ratio - increased in svPPA, #nouns/#words - reduced in svPPA, #verbs/#words - reduced in nfvPPA, proportion verbs - reduced in svPPA, word frequency - increased in svPPA, adverbs - increased in svPPA, brunets index - reduced in svPPA, words used once - reduced in svPPA, type token ratio - reduced in svPPA, mean length of clause - reduced in nfvPPA, clauses - reduced in nfvPPA, complex nominals per clause - reduced in nfvPPA and svPPA, T-units - increased in nfvPPA, mean length of T-unit - reduced in nfvPPA, complex nominals per T-unit - reduced in nfvPPA and svPPA, total Yngve depth - reduced in nfvPPA, max and mean Yngve depth - reduced in nfvPPA and svPPA |
| 2017 | Cordella | English | 8 HCs, 11 nfvPPA, 14 lvPPA, 13 svPPA | Picture description - picnic | total syllables/total response, articulation rate, pause frequency, proportion pause, mean pause duration | total syllables/total response - reduced in nfvPPA, lvPPA, and svPPA, articulation rate - reduced in nfvPPA, lvPPA, and svPPA, pause frequency - increased in nfvPPA, proportion pause - increased in nfvPPA and lvPPA, mean pause duration - increased in nfvPPA. |
| 2014 | Maruta | English | 17 HCs, 15 nfvPPA | reading task with DAF paradigm - grandfather passage | mean number of words/second, mean number of errors (speech sound errors/grammatical errors) | mean words/minute - reduced in nfvPPA. |
| 2012 | Sajjadi | English | 30 HCs, 16 SD | semi-structured interview, picture description | speech rate, unit length, spontaneity, combined phonological errors, fluency, discourse markers, hesitation markers, editing breaks, open- and closed-class words, Number of verbs per unit, semantic errors, circumlocutions, information content, pictorial theme, complex units, number of arguments, number of clauses, Open- and closed-class word errors, verb agreement errors. | Speech rate - decreased in SD, unit length - decreased in SD, strings of phonemes - increased in SD, hesitation markers - increased in SD, complete units - decreased in SD, elliptical phrases - increased in SD, abandoned utterances - increased in SD, discourse markers - increased in svPPA, editing breaks - increased in SD, semantic errors - increased in SD, closed-class words - increased in SD, circumlocutions - increased in SD, redundant words and phrases - increased in SD, pictorial themes - decreased in SD, complex units - decreased in SD, arguments per verb - decreased in SD, verb agreement errors - increased in SD. |
| 2010 | Wilson | English | 10 HCs, 14 nfvPPA, 25 svPPA, 11 lvPPA | elicited speech production task | false starts, repaired sequences, number of pauses, speech rate, maximum speech rate, total speaking time, total words, mean length utterance (MLU), words in sentences, proportion pronouns, verb tokens/word, proportion closed class words, verb frequency, nouns with determiners, embeddings phw, auxiliary complexity, phonological errors, syntactic errors, semantic errors, incomplete sentences, distortions per 100 words | repaired sequences - increased for svPPA, number of pauses - increased for nfvPPA, speech rate - decreased for nfvPPA, lvPPA, svPPA, maximum speech rate - decreased for nfvPPA, lvPPA, total speaking time - decreased for nfvPPA, lvPPA, bvFTD, total words - decreased for nfvPPA, mean length utterance (MLU) - decreased for nfvPPA, lvPPA, svPPA, words in sentences - increased for lvPPA, svPPA, proportion pronouns - decreased for nfvPPA, svPPA, verb tokens/word - increased for lvPPA, verb frequency - increased for svPPA, nouns with determiners - decreased for nfvPPA, syntactic errors - increased for lvPPA, incomplete sentences - increased for svPPA, distortions per 100 words -increased for nfvPPA. |
| 2010 | Suh | Korean | 134 HCs, 6 SD | Hanja and Hangul reading/writing tasks | Hanja and Hangul reading performance | Hanja reading performance - reduced in SDm |
| 2010 | Gross | English | 8 HCs, 20 CBS | narrative task – frog where are you | speech rate (WPM), narrative theme (participants’ ability to maintain the story theme), global connectedness (whether subjects identified the point of the story), lexical connectedness (conceptual connectedness between consecutive story events) | speech rate (WPM) – reduced in CBS, narrative theme – reduced in CBS, global connectedness – reduced in CBS, lexical connectedness – reduced in CBS |
| 2008 | Sachin | Hindi | 37 HCs, 12 PSP | semi-structured interview, reading task | maximum phonation time (MPT), semantic fluency and reading speed. | Maximum phonation time (MPT) – reduced in PSP, semantic fluency and reading speed – reduced in PSP |
| 2022 | Bruffaerts | Dutch | 24 HCs, 12 nfvPPA, 11 svPPA | picture description – cookie theft, repetition task – Akense Afasie task | PVI of vowel nucleus duration, vowel duration | PVI (weak-strong words) – reduced in nfvPPA, vowel duration (strong-weak words first vowel) – increased in nfvPPA, vowel duration (strong-weak words second vowel) – increased in nfvPPA, vowel duration (weak-strong words first vowel) – increased in nfvPPA |
| 2022 | Matias-Guiu | Spanish | 31 HCs, 31 nfvPPA, 45 lvPPA, 11 svPPA | picture description – cookie theft | speech rate (WPM), total words, nouns (token), verbs (token), percentage of verbs, number of open-class words, number of closed-class words, noun: verb ratio, nouns frequency, verb frequency, nouns per 100 words, percentage of mazes, number of different words, different words per time, number of errors, number of errors/number of words | speech rate – decreased in nfvPPA, lvPPA, total words – decreased in nfvPPA, insignificant in lvPPA, svPPA, nouns (token) – decreased in nfvPPA, lvPPA, svPPA, verbs (token) – decreased in nfvPPA, number of open-class words – increased in nfvPPA, lvPPA, svPPA, number of closed-class words – decreased in nfvPPA, lvPPA, nouns per 100 words – decreased for lvPPA, svPPA, percentage of mazes – increased in nfvPPA, lvPPA, svPPA, number of different words – decreased in nfvPPA, lvPPA, svPPA, different words per time – decreased in nfvPPA, lvPPA, svPPA, number of errors/number of words – increased in nfvPPA. |
| 2021 | Gallee | English | 31 HCs, 25 HCs, 26 lvPPA, 15 svPPA | Picture description - picnic | content units (raw and normalized), self-referential utterances, inability to express, tangential utterances, empty utterances, false starts | content units - decreased in nfvPPA, lvPPA, and svPPA, content units/total utterances - decreased in lvPPA, and svPPA, self-referential utterances - increased in svPPA, inability to express - increased in svPPA and lvPPA, empty utterances - increased in lvPPA, false starts - increased in lvPPA |
| 2021 | Mack | English | 40 HCs, 34 nfvPPA, 16 lvPPA, 15 svPPA, 12 mxPPA | Narrative task - Cinderella story, Sentence Production Priming Test (SPPT) and the Northwestern Anagram Test (NAT) | clause diversity, narrative accuracy | clause diversity - reduced for mxPPA, nfvPPA, lvPPA, svPPA, narrative accuracy - reduced for mxPPA, nfvPPA, lvPPA, svPPA |
| 2020 | Zimmerer | English | 20 HCs, 34 nfvPPA, 25 lvPPA, 29 svPPA | holiday narrative | total words, combination ratio (how often longer combinations are produced), content word count, content word frequency, collocation strength | total words - reduced in nfvPPA, bvFTD, content word frequency - increased for lvPPA, svPPA, bvFTD, connectedness - reduced for nfvPPA, lvPPA, bvFTD, collocations - increased for svPPA. |
| 2020 | Matias-Guiu | Spanish | 31 HCs, 23 nfvPPA, 41 lvPPA, | Narrative task - “Historia de un Pajarito” (A little bird story) | false starts, proportion pause, total pause duration, number of pauses, total reading time, total speaking time, percentage of speech, mean length utterances (MLU), mean duration utterances, time after full stops, sentence duration, pause number, pause duration, final rise, conduites d'approche, number of errors, phonological errors, number of errors short declarative, number of errors, function word changes | false starts - increased for nfvPPA, proportion pause - increased for nfvPPA, lvPPA, total pause duration - increased for nfvPPA, lvPPA, number of pauses - increased for nfvPPA, lvPPA, total reading time - decreased for nfvPPA, lvPPA, total speaking time - decreased for nfvPPA, lvPPA, percentage of speech - decreased for nfvPPA, lvPPA, mean length utterances (MLU) - decreased for nfvPPA, lvPPA, mean duration utterances - decreased for nfvPPA, lvPPA, time after full stops - increased for nfvPPA, sentence duration - increased for nfvPPA, lvPPA, pause number - increased for nfvPPA, lvPPA, pause duration - increased for nfvPPA, lvPPA, final rise - decreased for nfvPPA, conduites d'approche - increased for nfvPPA, lvPPA, phonological errors - increased for nfvPPA, lvPPA, number of errors short declarative - increased for nfvPPA, number of errors long declarative - increased for lvPPA, number of errors short exclamatory - increased for lvPPA, number of errors long exclamatory - increased for nfvPPA, lvPPA, number of errors short interrogative - increased for nfvPPA, number of errors long declarative - increased for nfvPPA, function word changes - increased for nfvPPA |
| 2019 | Ash | English | 36 HCs, 9 nfvPPA, 14 lvPPA, 11 svPPA | picture description - cookie theft | words per minute, speech errors/100 words, dependent clauses per 100 utterances, percentage of utterances that are well formed sentences | speech rate (WPM) - reduced for bvFTD, lvPPA, svPPA, and nfvPPA, speech errors per 100 words - reduced for lvPPA, svPPA, and nfvPPA, dependent clauses per 100 utterances - reduced for bvFTD, svPPA, and nfvPPA, percentage of utterances that are well formed sentences - reduced for bvFTD, lvPPA, svPPA, and nfvPPA |
| 2017 | Cousins | English | 31 HCs, 20 svPPA, 42 bvFTD | picture description - cookie theft | noun concreteness, number of words produced, number of nouns per 100 words | number of words produced - reduced in svPPA and bvFTD, number of nouns per 100 words - reduced in svPPA and bvFTD, noun concreteness - reduced in svPPA and bvFTD |
| 2017 | Staiger | German | 130 HCs, 17 PSP | reading task | articulation rate | articulation rate - reduced in PSP |
| 2016 | Ash | English | 27 HCs, 19 svPPA, 54 bvFTD, 33 CBS | picture description - cookie theft | quantifiers per 100 words, nouns per 100 words | quantifiers - reduced for bvFTD and CBS, insignificant for svPPA, nouns - reduced for svPPA |
| 2015 | Mack | English | 12 HCs, 12 nfvPPA, 12 svPPA, 11 lvPPA | Narrative task - Cinderella story | speech rate (WPM), mean length utterance (MLU), mean length phonemes, mean length phoneme nouns, nouns (token), verbs (token), open: closed class ratio, noun: verb ratio, verb frequency, noun frequency, %grammatical sentences | speech rate (WPM) - reduced for nfvPPA, mean length utterance (MLU) - reduced for nfvPPA, mean length phoneme nouns - reduced for nfvPPA, nouns (token) - reduced for nfvPPA, lvPPA, svPPA, verbs (token) - reduced for nfvPPA, open: closed class ratio - reduced for svPPA, noun: verb ratio insignificant for nfvPPA, lvPPA, svPPA, %grammatical sentences - reduced for nfvPPA, svPPA. |
| 1997 | Thompson | English | 5 HCs, 4 nfvPPA | narrative task - Cinderella story, picture description task - cookie theft, interview | mean length utterance (MLU), open: closed-class ratio, noun: verb ratio, %verbs with correct verb arguments, % complex units, proportion grammatical sentences | mean length utterance (MLU) - reduced in nfvPPA, open: closed-class ratio - increased in nfvPPA, noun: verb ratio - insignificant in nfvPPA, %verbs with correct verb arguments - reduced in nfvPPA, % complex units - reduced in nfvPPA, proportion grammatical sentences - reduced in nfvPPA |
| 2014 | Wilson | English | 13 HCs, 12 nfvPPA, 23 svPPA, 13 lvPPA | elicited speech production task | inflectional morphology | Real word inflection - reduced for nfvPPA, lvPPA, and svPPA, non-word inflection - reduced for nfvPPA, lvPPA. |
| 2014 | Silveri | Italian | 10 HCs, 21 nfvPPA, 11 svPPA, 3 lvPPA, 3 mxPPA | picture description - cookie theft and picnic description | mean length of utterance (MLU), speech rate (WPM) and percentage of fragments | speech rate - reduced in nfvPPA, lvPPA, svPPA, mean length utterance (MLU) - reduced in nfvPPA, lvPPA, svPPA |
| 2014 | Meteyard | English | 8 HCs, 8 SD | Semi-structured interview - Autobiographical Memory Interview | open class items, closed class items, noun and verb tokens, light verbs, light nouns, demonstrative pronouns, interrogative pronouns and indeterminate locative terms, open: closed class, light nouns: all nouns, light verbs: all verbs, word-form frequency analysis, Frequency of complex verb morphology, frequency of syntactic constructions | frequency - increased for SD, noun frequency - increased for SD, demonstrative pronouns - increased for SD, interrogative pronouns - increased for SD |
| 2014 | Hoffman | English | 8 HCs, 7 SD | Semi-structured interview - Autobiographical Memory Interview | noun frequency, imageability | noun frequency - increased in SD, imageability - decreased in SD |
| 2012 | DeLeon | English | 12 HCs, 16 nfvPPA, 7 svPPA, 8 lvPPA, 6 bvFTD | elicited speech production task | targeted structures attempted, attempted structures correct, syntactic errors, semantic errors | targeted structures attempted - decreased for nfvPPA, lvPPA, svPPA, attempted structures correct - decreased for nfvPPA, bvFTD, syntactic errors - increased for lvPPA, semantic errors - increased for svPPA |
| 2009 | Ash | English | 10 HCs, 11 nfvPPA, 12 SD, 12 bvFTD | Narrative task - Frog where are you | speech rate, total words, mean segment duration, mean length utterance (MLU), nouns (token), verbs (token) - decreased in nfvPPA, existential subjects, complex structures per utterance, errors/utterance | speech rate - decreased in nfvPPA, SD, bvFTD, total words - decreased in nfvPPA, bvFTD, mean length utterance (MLU) - decreased in nfvPPA, nouns (token) - decreased in nfvPPA, SD, verbs (token) - decreased in nfvPPA, existential subjects - increased in SD, complex structures per utterance - decreased in nfvPPA, errors/utterance - increased in nfvPPA |
| 2005 | Ash | English | 6 HCs, 8 nfvPPA, 9 svPPA | Narrative task - Frog where are you | WPM, MLU, #words, well-formed sentences, phrasal adjuncts, dependent clauses, %open class words, semantically deviant utterances, nouns/inflected words, errors on words | speech rate (WPM) - decreased in nfvPPA and svPPA, mean length utterance (MLU) - decreased in nfvPPA, proportion open-class words - decreased svPPA, ratio of nouns to inflected verbs - decreased nfvPPA, phrasal adjuncts - decreased nfvPPA, semantically deviant utterances - increased svPPA, well-formed sentences - decreased nfvPPA, dependent clauses per 100 utterances - decreased nfvPPA, errors on verbs - increased nfvPPA, svPPA |
| 2000 | Bird | English | 20 HCs, 3 SD | picture description - cookie theft | speech rate (WPM), ratio of content and function words, noun verb frequency, | content words: function words ratio - reduced in SD, verb frequency - increased in SD, noun frequency - increased in SD, |
| 2022 | Karpathiou | Greek | 15 HCs, 10 PPA | cookie theft and a story retell task | Articulation rate, speech rate, mean pause duration, number of pauses, total words, mean length sentences, sentence elaboration index, type token ratio, embedding index, narrative words/total words, mean log frequency of narrative words, dysfluencies per total words | Articulation rate - decreased in mxPPA, mean pause duration - increased in mxPPA, number of pauses - increased in mxPPA, total words - decreased in mxPPA, mean length sentences - decreased in mxPPA, sentence elaboration index - decreased in mxPPA, type token ratio - decreased in mxPPA, embedding index - decreased in mxPPA, narrative words/total words - decreased in mxPPA, mean log frequency of narrative words - increased in mxPPA, dysfluencies per total words - increased in mxPPA |
| 2022 | Rezaii | English | 53 HCs, 29 nfvPPA, 26 lvPPA, 24 svPPA | picture description - picnic scene, topic-directed interview | content word frequency, frequency of syntactic constructions | content word frequency - decreased in nfvPPA, increased in lvPPA, svPPA, frequency of syntactic constructions - increased in nfvPPA, decreased in lvPPA, svPPA |
| 2021 | Lavoie | English | 13 HCs, 13 lvPPA | picture description - cookie theft, narrative task - Cinderella, semi-structured interview | mean length of sample, proportion open-class words, proportion nouns, proportion pronouns, proportion verbs, proportion well-formed sentences | mean length of samples - insignificant for lvPPA, proportion of open-class words - reduced in lvPPA, proportion of verbs - increased in lvPPA, proportion of pronouns - increased in lvPPA, well-formed sentences - reduced in lvPPA |
| 2021 | Bouvier | Quebec French | 30 HCs, 9 PPAOS, 4 DAOS | repetition task - TEst Français de RÉpétition de Phrases (TEFREP) | Length of speech runs (number of syllables), Articulation rate (syllables/s), normalized pairwise variability index (nPVI) | Length of speech runs (number of syllables) - reduced in PPAOS, articulation rate (syllables/s) - reduced in PPAOS, normalized pairwise variability index (nPVI) - reduced in PPAOS |
| 2018 | Koukoulioti | Greek | 7 HCs, 7 SD | elicited speech production task | verb retrieval | verb production - decreased in SD |
| 2016 | Yunusova | English | 33 HCs, 9 nfvPPA, 9 bvFTD | reading task - bamboo passage | proportion pause, mean pause duration, total pause duration, number of pauses, pause duration variability, speech rate, articulation rate, mean phrase duration, coefficient of variation phrase | proportion pause - increased for nfvPPA, bvFTD, total pause duration - increased for nfvPPA, bvFTD, ALS-FTD, number of pauses - increased for nfvPPA, bvFTD, ALS-FTD, pause duration variability - increased for bvFTD, ALS-FTD, insignificant for nfvPPA, speech rate - decreased for nfvPPA, bvFTD, ALS-FTD, articulation rate - decreased for nfvPPA, ALS-FTD, insignificant for bvFTD, mean phrase duration - decreased for nfvPPA, ALS-FTD, coefficient of variation phrase - increased for nfvPPA, ALS-FTD |
| 2014 | Ballard | English | 17 HCs, 20 nfvPPA, 21 lvPPA | reading task - grandfather passage | syllable segregation, PVI, silence duration, variability of silence duration | median PVI strong-weak - reduced in nfvPPA, median PVI weak-strong - reduced in nfvPPA, proportion silence time - increased in nfvPPA, lvPPA, variability of silence duration - increased in nfvPPA, lvPPA |
| 2010 | Garrard, | English | 21 HCs, 21 SD | picture description - cookie theft | word count, speech rate (WPM), noun:verb ratio, ratio of content words: function words, | noun: verb - reduced for SD, content to function word ratio - reduced for SD |
| 2016 | Irish | English | 10 HCs, 10 svPPA | Semi-structured interview | use of past tense | use of past tense - reduced in svPPA |
| 2004 | Graham | English | 11 HCs, 14 nfvPPA | picture description (spoken and written) - cookie theft | speech rate (WPM), word count, information units, inaccurate/irrelevant information, grammatical endings on words, nouns and verbs, content, and function verbs, paraphasia’s and neologisms | paraphasia’s/words - increased in nfvPPA, paraphasia’s - increased in nfvPPA, speech rate (WPM) - decreased in nfvPPA, total speaking time - decreased in nfvPPA, total words - decreased in nfvPPA, nouns (token) - decreased in nfvPPA, verbs (token) - decreased in nfvPPA, nouns (type) - decreased in nfvPPA, verbs (type) - decreased in nfvPPA, #verbs/#words - decreased in nfvPPA, content words as proportion of total words - decreased in nfvPPA, function words as proportion of total words - decreased in nfvPPA, neologisms - increased in nfvPPA, neologisms/word - increased in nfvPPA, content units - decreased in nfvPPA, grammatical errors - increased in nfvPPA |
| 2009 | Meteyard | English | 8 HCs, 8 SD | semi-structured interview | total words, mean length utterance, insertions, word omissions, phonological errors, substitution errors, perseveration/anticipation errors, complex morphological errors, simple morphological errors, planning errors, open-class errors, closed-class errors | total words - decreased in SD, mean length utterance (MLU) - decreased in SD, insertions - insignificant in SD, word omissions - increased in SD, phonological errors - insignificant in SD, substitution errors - increased in SD, perseveration/anticipation errors - increased in SD, complex morphological errors - increased in SD, simple morphological errors - insignificant in SD, planning errors - increased in SD, open-class errors - increased in SD, closed-class errors - increased in SD |
| 2015 | Rusz | Czech | 37 HCs, 12 PSP | 90s monologue | % Pause time, no. pauses, mean speech intensity, pitch variability, vowel articulation index, percentage dysfluent words, articulation rate, intensity variations. | intensity - decreased in PSP, intensity variability - increased in PSP, percentage pause - increased in PSP, articulation rate - decreased in PSP, vowel articulation index, % dysfluent words - increased in PSP |
| 2020 | Jarrold | English | 10 HCs, 14 nfvPPA, 25 svPPA, 11 lvPPA, bvFTD | Picture description - picnic | ratio of pronouns to verbs | pronoun to verb ratio - increased for svPPA |
| 2019 | Cordella | English | 20 HCs, 22 nfvPPA, 23 lvPPA, 19 svPPA | Picture description - picnic | Articulation rate (syllables per second) | articulation rate - reduced for nfvPPA, lvPPA, and svPPA |
| 2010 | Ash | English | 10 HCs, 16 nfvPPA | Narrative task - Frog where are you | total words, speech rate (WPM), mean length utterance (MLU), editing breaks/hesitation markers per 100 words, open class words per utterance, nouns per utterance, total verbs per utterance, complex structures per utterance, proportion of well-formed utterances | editing breaks - increased in nfvPPA, hesitation markers - increased in nfvPPA, speech rate - decreased in nfvPPA, total words - decreased in nfvPPA, mean length utterance (MLU) - decreased in nfvPPA, verbs per utterance - decreased in nfvPPA, open class words per utterance - decreased in nfvPPA, nouns per utterance - decreased in nfvPPA, complex structures per utterance - decreased in nfvPPA, well-formed sentences - decreased in nfvPPA |
| 2016 | Rogalski | English | 35 HCs, 6 nfvPPA, 13 lvPPA | Narrative task | speech rate (WPM) | speech rate (WPM) - decreased in nfvPPA and lvPPA |
| 2019 | Berube | English | 50 HCs, 44 PPA | Picture description task - updated cookie theft | number of syllables, content units (CUs), syllables per CU, and the ratio of left-right CUs (information from left and right sides of image) | number of syllables - reduced in mxPPA, content units (CUs) - reduced in mxPPA, syllables per CU - reduced in mxPPA, ratio of left-right CUs - increased in mxPPA |
| 2019 | Catricala | Italian | 27 HCs, 17 PSP | Picture description task - seaside scene | speech rate (total words/total time), total locution time, number of pauses (number/total locution time), between-utterance pause duration, phonemic errors (Well-articulated phoneme substitutions, additions, transpositions, and deletions/total words), noun rate, verb rate, pronoun rate, noun-verb ratio, quantifiers, repaired sentences, semantic errors, mean length sentences, total sentences, incomplete sentences, dependent clauses, morpho-syntactic errors, total words, information units, micro proposition, implausible or irrelevant details, index of discourse effectiveness (The ratio of the total number of recalled words divided by the number of information units. Index of discourse effectiveness/total sentences), errors in content elements, referential cohesion errors (Total number of referential cohesive ties (pronouns), used in an ambiguous or erroneous way. Referential cohesion errors/total pronouns), efficiency | percentage of speech - decreased for PSP, syllable duration variability - decreased for PSP, number of sentences - decreased for PSP, mean length of sentence - decreased for PSP, proportion pronouns - increased for PSP, maintenance of search theme - decreased for PSP, incomplete sentences - increased for PSP |
| 2016 | Hardy | English | 24 HCs, 18 nfvPPA, 14 svPPA, 24 bvFTD | holiday narrative | total words, mean words/prompt, words frequency | Total words - reduced in nfvPPA, mean words per prompt - reduced for bvFTD, nfvPPA, svPPA, word frequency - increased for nfvPPA and svPPA |
| 2012 | Sajjadi | English | 30 HCs, 12 nfvPPA, 16 mxPPA | semi-structured interview, picture description task - man in armchair | speech rate, unit length, frequency of elliptical phrases, frequency of discourse markers, frequency of hesitation markers, frequency of editing breaks, frequency of relative sentences, closed class insertions, open class omission errors, verb agreement errors, speech rate, number of discrete speech units/100 words, arguments per verb, number of clauses, closed-class errors, frequency of closed-class omissions, verb agreement errors | speech rate - reduced in nfvPPA and mxPPA, unit length - reduced in nfvPPA and mxPPA, number of discrete speech units/100 words - reduced in nfvPPA and mxPPA, speech sound errors - increased in nfvPPA and mxPPA, hesitation markers - increased in nfvPPA and mxPPA, production of compete units - reduced in nfvPPA and mxPPA, elliptical phrases - increased in nfvPPA and mxPPA, abandoned units - increased in mxPPA, editing breaks - increased in nfvPPA and mxPPA, discourse markers - increased in mxPPA, closed class words - increased in mxPPA, semantic errors - increased in mxPPA, open-class words errors - increased in nfvPPA, redundant words and phrases - increased in mxPPA and nfvPPA, mentioned pictorial themes in picture description - decreased in mxPPA and nfvPPA, complex units - reduced in nfvPPA and mxPPA, arguments per verb - reduced in mxPPA, number of clauses - insignificant for nfvPPA and mxPPA, closed-class errors - increased in nfvPPA, verb agreement errors - increased in mxPPA |
| 2010 | Pakhomov | English | 32 HCs, 12 nfvPPA, 11 svPPA, 6 lvPPA, 19 bvFTD | picture description - cookie theft | perplexity index (degree of deviation in word patterns used), out of vocabulary rate (proportion of words used by FTLD patients that were not used by HCs) | out of vocabulary rate - increased for svPPA |
| 2020 | Faroqi-Shah | English | 25 HCs, 10 nfvPPA, 9 lvPPA, 7 svPPA | picture description - cookie theft | WPM, total number of dysfluencies, moving average type token ratio, idea density (# of propositions (non-noun entities)/ 10 words), CUs (Total # correct information units/ Total # words in sample), total semantic errors, total phonological errors, total word retrieval errors, MLU, proportion of grammatical utterances, verbs per utterance, sentence complexity, total morphological errors, total errors | speech rate (WPM) - reduced in PPA, total number of dysfluencies - increased in PPA, CUs (Total # correct information units/ Total # words in sample) - reduced in PPA, total phonological errors- increased in PPA, total word retrieval errors - increased in PPA, proportion of grammatical utterances - reduced in PPA, total errors - increased in PPA. |
| 2014 | Marcotte | English | 15 HCs, 12 nfvPPA, 11 svPPA | story completion task | verb retrieval, errors in heavy/light verbs, grammaticality | grammatically correct responses - decreased in nfvPPA, errors in general verbs - increased in nfvPPA, errors in specific verbs - increased in nfvPPA, svPPA, errors in heavy verbs - increased in nfvPPA, errors in light verbs - increased in nfvPPA |
| 2013 | Ash | English | 12 HCs, 15 nfvPPA, 29 lvPPA, 18 svPPA, 17 bvFTD | picture description - cookie theft | speech rate (WPM), total words, MLU, percentage open-class words, nouns per 100 words, well-formed sentences, dependent clauses per 100 utterances, phonetic errors, phonemic errors, fluency disruptions per 100 words | speech rate (WPM) - decreased in nfvPPA, lvPPA, svPPA, total words - decreased in nfvPPA, lvPPA, svPPA, bvFTD, MLU - decreased in nfvPPA, percentage open-class words - decreased in lvPPA, svPPA, nouns per 100 words - decreased in svPPA, well-formed sentences - decreased in lvPPA, svPPA, dependent clauses per 100 utterances - decreased in nfvPPA, phonemic errors - increased in nfvPPA, fluency disruptions per 100 words - increased in lvPPA |
| 2016 | Santos-Santos | English | 10 HCs, 5 PSP, 9 CBS | Picture description - picnic | total narrative words, speech rate (WPM), proportion of syntactic errors, proportion of words in sentences, proportion of distortions (per 100 words) | speech rate (WPM) - decreased in nfvPPA, total words - decreased in nfvPPA, proportion words in sentences - decreased in nfvPPA, syntactic errors - increased in nfvPPA, distortions/100 words - increased in nfvPPA |
| 2014 | Fraser | English | 16 HCs, 14 nfvPPA, 10 SD | Narrative task - Cinderella story | total words/total time, total words, mean length sentences, mean length clause, mean length clause, word frequency, number of sentences, mean word length, pronoun ratio, #nouns/#nouns + #verbs, nouns (token), verbs (token), noun: verb ratio, word frequency, verb frequency, noun frequency, imageability, noun imageability, verb imageability, light verbs, demonstratives, adverbs, adjectives, determiners, prepositions, type-token ratio, real word inflection, clauses, clauses per sentence, clauses per t-unit, complex t-units per t-unit, coordinate phrases per t-unit, complex nominals per t-unit, t-units per sentence, coordinate phrases per clauses, tree height, t-units, complex t-units, complex nominals, complex nominals per t-unit, total Yngve depth, max Yngve depth, mean Yngve depth, verb phrases, verb phrases per t-unit, dependent clauses per clauses, total dependent clauses, dependent clauses per t-unit, function words, fillers, age of acquisition (AOA), noun AOA, verb AOA, familiarity, noun familiarity, verb familiarity, coordinate phrases, number of um/uh's | total words/total time - reduced in nfvPPA and SD, total words - reduced in nfvPPA, mean length sentences - reduced in SD, mean length clause - reduced in SD, word frequency - increased for SD and nfvPPA, mean word length - reduced in nfvPPA and SD, pronoun ratio - increased for SD, #nouns/#nouns + #verbs - reduced in SD, nouns (token) - reduced in SD, noun: verb ratio - reduced in SD, word frequency - increased for SD and nfvPPA, verb frequency - increased for SD and nfvPPA, noun frequency - increased in SD, verb imageability - increased for SD, demonstratives - increased in nfvPPA and SD, adverbs - increased in SD, type-token ratio - decreased in SD, clauses - increased in SD, t-units per sentence - reduced in in nfvPPA and SD, complex nominals per t-unit - reduced for nfvPPA, mean Yngve depth - reduced in SD, age of acquisition (AOA) - reduced in nfvPPA noun AOA - reduced in nfvPPA, familiarity - increased in SD, noun familiarity - increased in SD, verb familiarity - decreased in SD |
| 2012 | Thompson | English | 13 HCs, 11 nfvPPA, 20 lvPPA, 6 svPPA | Narrative task - Cinderella story | speech rate (WPM), mean length of utterance in words (MLU-W), proportion of grammatically correct sentences, open-to-closed-class word ratio, noun-to-verb ratio, correct production of verb inflection, noun morphology, and verb argument structure | speech rate (WPM) - reduced in nfvPPA and lvPPA, mean length utterance (MLU) - reduced in nfvPPA and lvPPA, proportion of grammatically correct sentences - reduced in nfvPPA and lvPPA, open-to-closed-class word ratio - reduced in svPPA, noun-to-verb ratio - reduced in svPPA, production of verb inflection - reduced in nfvPPA, verb argument structure - reduced in nfvPPA |
| 2022 | Baque | Spanish | 4 HCs, 4 nfvPPA, 2 svPPA, 4 lvPPA, 5 bvFTD 4 ALS-FTD | reading task | total silence duration, median/mean silence length, variability of silence duration, silent pause: filled pause + speech, silent pause: total speech duration, articulation rate, total reading time, percentage of speech, total words/total time, speech segments between pauses, sum of speech segments between pauses, duration of longest phonic group, mean duration of phonic groups between silent pauses, median duration of phonic groups, standard deviation of the duration of the phonic groups, total number of syllabic nuclei of the longest phonic group obtained manually, mean duration of the syllables of the phonic group of maximum duration | articulation rate - decreased for nfvPPA, total reading time - decreased for lvPPA, speech segments between pauses - increased for nfvPPA, lvPPA, mean duration of the syllables of the phonic group of maximum duration - increased for lvPPA |
| 2009 | Knibb | English | HCs 15 HCs, 15 nfvPPA | semi-structured interview | speech rate (WPM), unit length, noun: verb ratio, elliptical phrases, subordinate syntactic units, grammatical errors, speech sound errors, closed-class errors | speech rate (WPM) - reduced in nfvPPA, unit length - reduced in nfvPPA, elliptical phrases - increased in nfvPPA, subordinate syntactic units - reduced in nfvPPA, closed-class errors - increased in nfvPPA |
| 2010 | Rohrer | English | 18 HCs, 24 nfvPPA | picture description - cookie theft | speech rate (WPM), agrammatic errors/min, speech production errors/min, mean pause length, frequency of nouns, frequency of verbs used | speech rate (WPM) - decreased for nfvPPA, agrammatic errors/min - increased for nfvPPA, production errors/min - increased for nfvPPA, mean pause length - decreased for nfvPPA, frequency of nouns - increased for svPPA, frequency of verbs used - increased for nfvPPA |
| 2006 | Ash | English | 10 HCs, 10 nfvPPA, 13 SD, 12 bvFTD (SOC/EXEC) | Narrative task - Frog where are you | duration of narrative sample, number of utterances, number of words, lexical retrieval difficulty (1) occurrence of word-finding difficulty, 2) a general noun in place of a specific one, 3) use of a wrong noun, 4) use of a wrong verb, and 5) a pronoun missing its antecedent), action, global connectedness, search theme, local connectedness, | speech rate (WPM) - reduced for nfvPPA, SD, bvFTD (EXEC/SOC), mean length utterance -reduced for nfvPPA, impaired word finding frequency - reduced for SD, errors -increased for nfvPPA, SD, bvFTD (EXEC/SOC), global connectedness - reduced for bvFTD (EXEC/SOC), maintenance of search theme - reduced for nfvPPA and bvFTD (EXEC/SOC), local connectedness - reduced for nfvPPA and SD |
| 2009 | Mesulam | English | 17 HCs, 4 nfvPPA, 7 lvPPA, 5 svPPA | Picture description - picnic | phrase length | phrase length - reduced in nfvPPA |
| 2022 | Cho | English | 28 HCs, 21 lvPPA | picture description - cookie theft | Articulation rate, percentage of speech, total words, mean segment duration, word frequency, mean word length, nouns (token), noun concreteness, adverbs, prepositions, age of acquisition, word repetitions | Articulation rate - reduced in lvPPA, percentage of speech - reduced in lvPPA, total words - reduced in lvPPA, mean segment duration - reduced in lvPPA, word frequency - increased in lvPPA, mean word length - reduced in lvPPA, nouns (token) - reduced in lvPPA, noun concreteness - reduced in lvPPA, adverbs - increased in lvPPA, prepositions - reduced in lvPPA, age of acquisition - reduced in lvPPA, word repetitions - increased in lvPPA |
| 2006 | Patterson | English | 24 HCs, 14 SD | reading task, writing to dictation, inflecting verbs task | reading performance low frequency words, reading performance irregular words | reading performance low frequency words - reduced in SD, reading performance irregular words - reduced in SD |
| 2020 | Koukoulioti | Greek | 5 HCs, 5 SD | sentence completion task | inflection past perfect/ present, inflection present, inflection past perfect/ imperfect, inflection past imperfect | inflection past perfect/ present - increased in SD, inflection present - increased in SD, inflection past perfect/ imperfect - increased in SD, inflection past imperfect - increased in SD |
| 2011 | Pakhomov | English | 25 HCs, 12 nfvPPA, 5 lvPPA, 11 svPPA, 17 bvFTD | picture description - cookie theft | periodicity of speech - g-statistic, average mode of the power spectra, average r-index, principal radial frequency across the power spectra | average r-index - significant difference in the FTLD group, principal radial frequency across the power spectra - significant difference in the FTLD group |
